# Delayed booster dosing improves human antigen-specific Ig and B cell responses to the RH5.1/AS01_B_ malaria vaccine

**DOI:** 10.1101/2022.04.25.22274161

**Authors:** CM Nielsen, JR Barrett, C Davis, JK Fallon, C Goh, AR Michell, C Griffin, A Kwok, C Loos, S Darko, F Laboune, SE Silk, M Tekman, JR Francica, A Ransier, RO Payne, AM Minassian, DA Lauffenburger, RA Seder, DC Douek, G Alter, SJ Draper

## Abstract

**Background:** Antibodies are crucial for vaccine-mediated protection against many pathogens. Modifications to vaccine delivery that increase antibody magnitude, longevity, and/or quality are therefore of great interest for maximising efficacy. We have previously shown that a delayed fractional (DFx) dosing schedule (0-1-6mo) – using AS01_B_-adjuvanted RH5.1 malaria antigen – substantially improves serum IgG durability as compared to monthly dosing (0-1-2mo; NCT02927145). However, the underlying mechanism and whether there are wider immunological changes with DFx dosing was unclear.

**Methods:** Immunokinetics of PfRH5-specific Ig across multiple isotypes were compared between DFx and monthly regimen vaccinees. Peak responses were characterised in-depth with a systems serology platform including biophysical and functional profiling. Computational modelling was used to define the humoral feature set associated with DFx dosing. PfRH5-specific B cells were quantified by flow cytometry and sorted for single cell RNA sequencing (scRNA-seq). Differential gene expression between DFx and monthly dosing regimens was explored with Seurat, DESeq2 and gene set enrichment analysis.

**Results:** DFx dosing increases the frequency of circulating PfRH5-specific B cells and longevity of PfRH5-specific IgG1, as well as other isotypes and subclasses. At the peak antibody response, DFx dosing was distinguished by a systems serology feature set comprising increased FcRn-binding, IgG avidity, and proportion of G2B and G2S2F IgG Fc glycans, alongside decreased IgG3, antibody-dependent complement deposition, and proportion of G1S1F IgG Fc glycan. At the same time point, scRNA-seq of PfRH5-specific B cells revealed enriched plasma cell and Ig / protein export signals in the monthly dosing group as compared to DFx vaccinees.

**Conclusions:** DFx dosing of the RH5.1/AS01_B_ vaccine had a profound impact on the humoral response. Our data suggest plausible mechanisms relating to improved FcRn-binding (known to improve Ig longevity) and a potential shift from short-lived to long-lived plasma cells. Recent reports of the positive impact of delayed boosting on SARS-CoV-2 vaccine immunogenicity highlight the broad relevance of these data.

## Introduction

Vaccines are among public health’s most effective tools for combatting infectious disease but a poor understanding of the underlying immunological mechanisms frequently impedes vaccine development. One of the greatest perennial issues for the vaccinology field is a lack of knowledge of how to induce more *durable* immune responses in the target populations. While many vaccine candidates generate encouraging peak antibody concentrations, these often wane rapidly in the following months. This rapid decay can be highly problematic and poses a particular issue for pathogens such as blood-stage malaria where the threshold antibody concentration required for protection is high (1, 2).

Interestingly, in a clinical trial with the leading blood-stage malaria vaccine candidate (RH5.1/ AS01_B_) we have recently shown that a delayed fractional booster 0-1-6-month schedule (“DFx”) induces higher magnitude vaccine-specific antibody as compared to a more typically used 0-1-2-month “monthly” vaccination schedule (NCT02927145). Moreover, the DFx anti-PfRH5 serum IgG plateaus at a 10X higher concentration over the next 2 years – an unprecedented finding (1). While data from other malaria vaccine trials – and more recent SARS-CoV-2 trials (3–5) – are broadly supportive of a beneficial impact of delayed (fractional) booster dosing on antibody-mediated immunity, there is yet to be any other demonstration of a comparable impact of vaccine regimen on human antibody longevity or any analyses involving the direct detection and isolation of antigen-specific B cells (6–9).

Here, using samples from the RH5.1/AS01_B_ trial, we interrogate the PfRH5-specific antibody and B cell responses in both DFx and monthly regimens (**Table 1**). Through a combination of immunokinetic, systems serology, and single cell RNA sequencing (scRNA-seq) analyses, we identify features of the PfRH5-specific Ig and B cell responses that discriminate between these dosing regimens. These data are informative for understanding the potential underlying mechanisms of DFx-mediated improvements in humoral immunity, and will be of great relevance for efforts to further optimise durable antibody responses against malaria and other diseases where vaccine-induced protection is antibody-mediated. This work builds on previously published data demonstrating improved PfRH5-specific IgG and Tfh2 immunogenicity following PfRH5 delivery by RH5.1/AS01_B_ as compared to a heterologous viral vector platform (NCT02181088) (1, 10, 11).

**Table 1.** Overview of vaccination regimen. PfRH5.1 = full-length PfRH5 protein (1, 12). Vaccinee demographics relate only to those included in this study.

| Type of booster dosing regimen | VAC063 trial group | Vaccination time point and RH5.1 dose<br>(all vaccinations co-administered with AS01<sub>B</sub> adjuvant) |  |  |  | Vaccinee Demographics |  |
| --- | --- | --- | --- | --- | --- | --- | --- |
|  |  | Day 0 | Day 28 | Day 56 | Day 182 | % female | Median age, years (range) |
| Monthly | <sup>1</sup><br>'Monthly-low' | 2µg | 2µg | 2µg |  | 65<br>(22/34) | 31<br>(18-46) |
|  | <sup>2</sup><br>'Monthly-medium' | 10µg | 10µg | 10µg |  |  |  |
|  | <sup>4</sup><br>'Monthly-high' | 50µg | 50µg | 50µg |  |  |  |
| Delayed fractional | <sup>3</sup><br>'DFx' | 50µg | 50µg |  | 10µg | 67<br>(8/12) | 28<br>(25-42) |

## Results

### Delayed fractional (DFx) dosing improves longevity of circulating PfRH5-specific B cells and IgG1

Using PfRH5 probes to detect circulating PfRH5-specific memory IgG+ B cells (defined as live CD19+CD21+CD27+IgG+probe++ single lymphocytes; **Supplemental Figure 1A**), we first established that protein/AS01_B_ vaccinees had higher frequencies of these antigen-specific cells in circulation at both 4-weeks and 12-weeks after final vaccination as compared to heterologous viral vector vaccinees (**Figure 1A**). Within the protein/AS01_B_ trial, vaccinees receiving a DFx regimen (**Table 1**) – rather than the monthly dosing regimen – showed higher responses 2-, 4-, and 12-weeks after the final vaccination (**Figure 1B**). The discrepancy was even more pronounced if PfRH5-specific cells were defined less conservatively with only one probe: significant differences were sustained out to 12-weeks following final vaccination (PfRH5-PE; **Supplemental Figure 1B,C**). While the source of circulating anti-PfRH5 serum IgG is presumably bone marrow resident long-lived plasma cells (LLPCs) rather than circulating anti-PfRH5 memory B cells (mBCs), these two parameters do correlate at 4-weeks after the final vaccination suggesting a more robust B cell response with the DFx regimen as compared to monthly regimen across multiple germinal centre B cell fate lineages (**Figure 1C**). PfRH5-specific memory IgM+ responses were detectable but minimal (**Supplemental Figure 1D**). These results are also consistent with reports from similar “Fx017M” dosing (0-1-7mo) with the PfCSP-based pre-erythrocytic malaria vaccine RTS,S. In this instance, PfCSP-specific mBCs were indirectly detected at higher frequencies in Fx017M vaccinees as compared to the monthly 0-1-2mo regimen following 5-day PfCSP stimulation (9).

**Figure 1.**
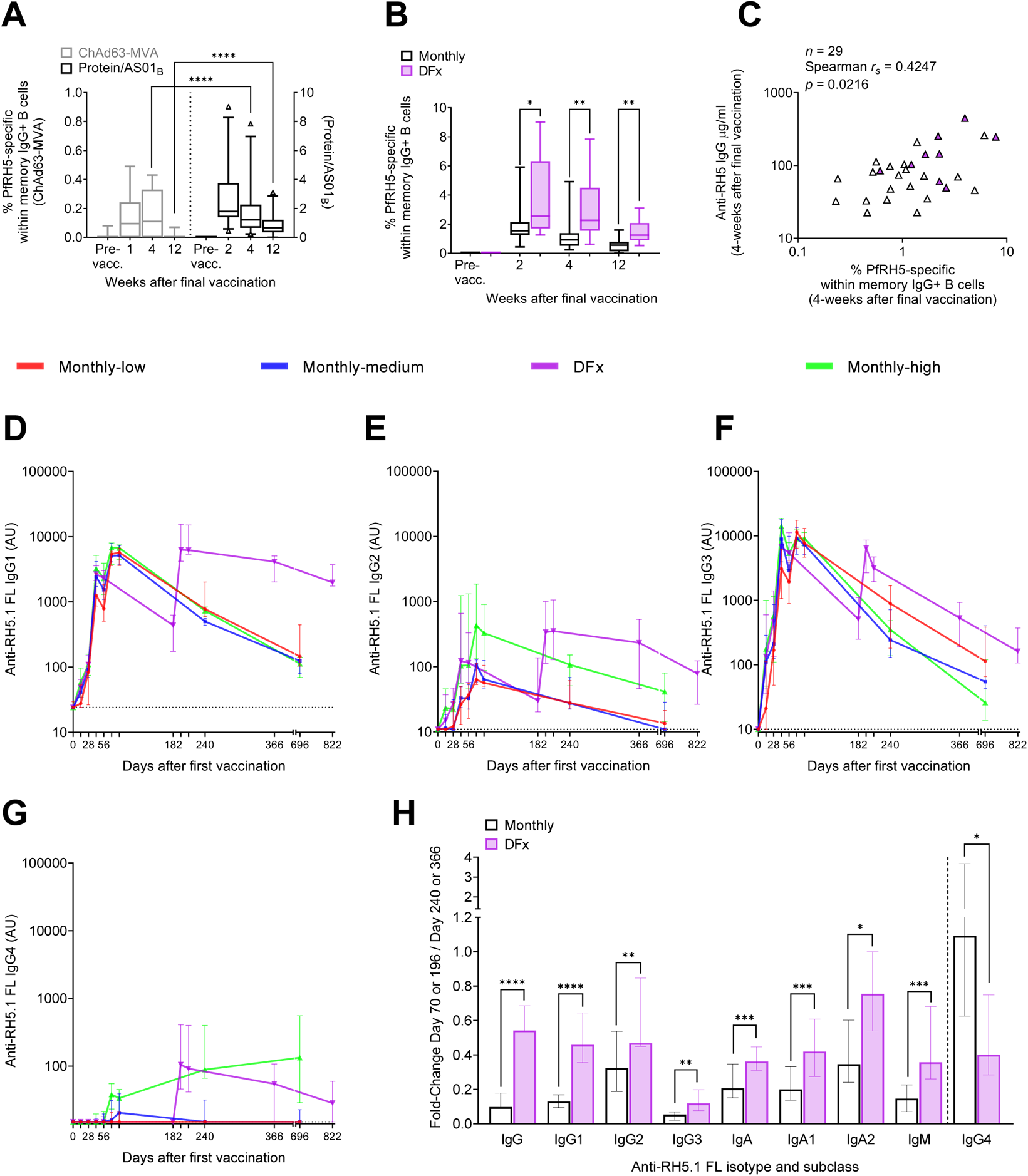
Antigen-specific B cell and Ig post-vaccination kinetics in DFx and monthly dosing regimens. PBMC from pre-vaccination (Pre-vacc.), and 1-, 2-, 4-, and 12-weeks post final vaccination time points were enriched for B cells and then stained with phenotypic markers and analysed by flow cytometry. Frequencies of PfRH5-specific mBCs were defined as CD19+CD21+CD27+IgG+IgM− cells within the live lymphocyte population that stained for monobiotinylated-PfRH5 conjugated to streptavidin-PE (gating strategy shown in **Supplemental Figure 1A**). Frequencies of PfRH5-specific mBCs were compared between (**A**) samples from a heterologous viral vector trial (ChAd63-MVA; ChAd63-PfRH5 prime, MVA-PfRH5 boost (10, 11) and the protein/AS01_B_ trial, or (**B**) between monthly regimen vaccinees and DFx vaccinees within the protein/AS01_B_ trial. (**C**) Spearman correlation analysis was performed between the frequency of PfRH5-specific mBCs 4-weeks after the final vaccination and the concentration of anti-PfRH5 serum IgG at the same time point (protein/AS01_B_ vaccinees only). Vaccinees were included in these PBMC analyses based on sample availability. (**A**) ChAd63-MVA/ protein/AS01B: Pre-vacc. *n* = 15/18; 1-week post final vaccination *n* = 10/0; 2-week *n* = 0/25; 4-week *n* = 15/29; 12-week *n* = 13/25. (**B**) Monthly / DFx within protein/AS01_B_ trial: Pre-vacc. *n* = 15/3; 2-week *n* = 16/9; 4-week *n* = 19/10; 12-week *n* = 17/8. (**A-B**) Comparisons were performed by Mann-Whitney tests. Whiskers denote 5^th^ and 95^th^ percentiles. * *p* < 0.05, ** *p* < 0.01, **** *p* < 0.0001. (**C**) Sample size, Spearman r, and p value are annotated on the graph. Each triangle represents one vaccinee and purple triangles indicate DFx vaccinees. Anti-RH5.1 FL (full length RH5.1) serum Ig was assayed by standardised ELISA to report (**D**) IgG1, (**E**) IgG2, (**F**) IgG3, and (**G**) IgG4 at key time points (Days 0, 14, 28, 42, 56/182, 70/196, 84/210, 240/366, 696/822). (**D-G**) Sample sizes for these ELISAs varied by group and by time point. Monthly-low: *n* = 12, except Day 696 where *n* = 9. Monthly-medium: *n* = 12, except for Day 240 and Day 696 where *n* = 11 and *n* = 10, respectively. DFx: *n* = 12, except for Day 366 and Day 822 where *n* = 11 and *n* = 7, respectively. Monthly-high: *n* = 11, except for Days 70, 240 and 696 where *n* = 9, *n* = 10 and *n* = 4, respectively. Graphs show medians and interquartile ranges. (**H**) Fold change in serum anti-PfRH5.1 FL Ig between 2-weeks after final vaccination (Day 70 for monthly regimen vaccinees, and Day 196 for DFx vaccinees) and 6-months after final vaccination (Day 240, Day 366). Monthly regimen/ DFx: *n* = 31/ 11 with the exception of IgG4 where *n* = 18 /9. Comparisons were performed between DFx and monthly regimens with Mann-Whitney tests. Bars denote medians and error bars denote interquartile ranges. * *p* < 0.05, ** *p* < 0.01, *** *p* < 0.001, **** *p* < 0.0001. Full kinetics for IgA, IgA1, IgA2 and IgM, as well a stratified version of panel H are shown in **Supplemental Figure 2**.

To delve further into the differences in anti-PfRH5 Ig immunokinetics between DFx and monthly vaccinees, we developed standardised ELISAs for anti-PfRH5 IgG1-4, IgA, IgA1-2, and IgM to assay sera samples from key post-vaccination time points (**Figure 1D-G**; **Supplemental Figure 2A-D**). Although there was no significant difference at the peak time points (2- or 4-weeks following final vaccination), 6-months after the final vaccination the median IgG1 AU was 4130 for DFx vaccinees as compared to 628AU in the monthly regimen vaccinees, i.e. ≈5-fold increase (*p*<0.0001; Mann Whitney test). After >1.5years, the IgG1 median AU values were 1984 and 133, respectively, i.e. ≈15-fold difference (*p*<0.0001; Mann Whitney test). In the absence of such stark differences in IgG2 (**Figure 1E**), IgG3 (**Figure 1F**) and IgG4 (**Figure 1G**), these data suggest that the majority of the improvement in longevity observed in the total anti-PfRH5-specific IgG response (10) is attributable to the IgG1 component. In fact, while levels of anti-PfRH5 IgG3 are not significantly different at 6-months or 1.5-years after the final vaccination (**Figure 1F**), both IgG2 and IgG4 show interesting indications that monthly-high vaccinees behave similarly to DFx, with trends towards high levels following the final vaccination (**Figures 1E, G**).

Given our interest in serum antibody maintenance over time – which may be modulated independently from changes in magnitude of the peak responses – we next calculated the fold change for each of the isotypes and subclasses between the peak response 2-weeks post final vaccination (Day 70 or Day 196) and the 6-month time point (Day 240 or 366), which had greater statistical power than the 1.5-year time point due to sample availability (**Figure 1H**). This clearly visualised the slower decay in total IgG, as reported previously (1), and IgG1 in the DFx vaccinees but also detected a significant difference in all other isotypes and subclasses between DFx vaccinees and (pooled) monthly regimen vaccinees – most convincingly for IgG3, IgA, IgA1 and IgM. This is an interesting indication that while DFx may not increase the magnitude of these responses, there may be an improvement in serum antibody response maintenance. Of note, IgG4 is the only isotype or subclass measured where decay is significantly faster with the DFx regimen – a signal which is largely driven by the unusual kinetics in the monthly-high vaccinees (**Figure 1G; Supplemental Figure 2E**).

### Systems serology feature set associated with DFx dosing includes increased FcRn-binding, IgG avidity, and proportion of bi-galactosylated IgG Fc glycans

Our stratified antibody isotype and subclass immunokinetic data suggest a significant impact of DFx dosing on humoral immunogenicity. We therefore next extended these analyses using a systems serology pipeline to integrate data on Fc biophysical and functional characteristics, measuring a total of 49 parameters (see Methods (13, 14)). As previously described (1), in addition to quantification of post-vaccination plasma levels of PfRH5-specific antibodies of each major isotype and subclass (with a Luminex bead-based assay rather than ELISA as above) this approach extends the biophysical analyses to include characterisation of the glycosylation profile of the anti-PfRH5 IgG Fc domains, which is known to influence these Fc-mediated functions (15). To assess Fc-mediated functionality of PfRH5-specific antibodies, the systems serology platform incorporates evaluation of their capacity to bind Fc receptors (FcRs), and to activate monocytes, neutrophils, natural killer (NK) cells, as well as the complement cascade. Here, in initial univariate analyses, we observed regimen-dependent differences in Fc-mediated activation with intriguing trends towards *reduced* functionality in DFx as compared to the monthly-low or monthly-medium vaccinees (**Figure 2**). Specifically, PfRH5-specific Ig in plasma samples 2-weeks following the final vaccination initiated decreased antibody-dependent complement deposition (ADCD; **Figure 2A**), antibody-dependent neutrophil phagocytosis (ADNP; **Figure 2B**) and NK cell cytokine production (MIP1β or IFNγ; **Figure 2C**). No differences were observed in NK cell degranulation as measured by CD107a expression (**Figure 2C**) or antibody-dependent cellular (monocyte) phagocytosis (ADCP; **Figure 2D**). It was of interest to note that in each instance samples from monthly-high vaccinees performed comparably to DFx vaccinees. This suggested that there was possibly an impact of the ‘high’ 50µg first and second doses (shared by these two groups) on Fc-mediated functionality, independent from any effect of a DFx final booster.

**Figure 2.**
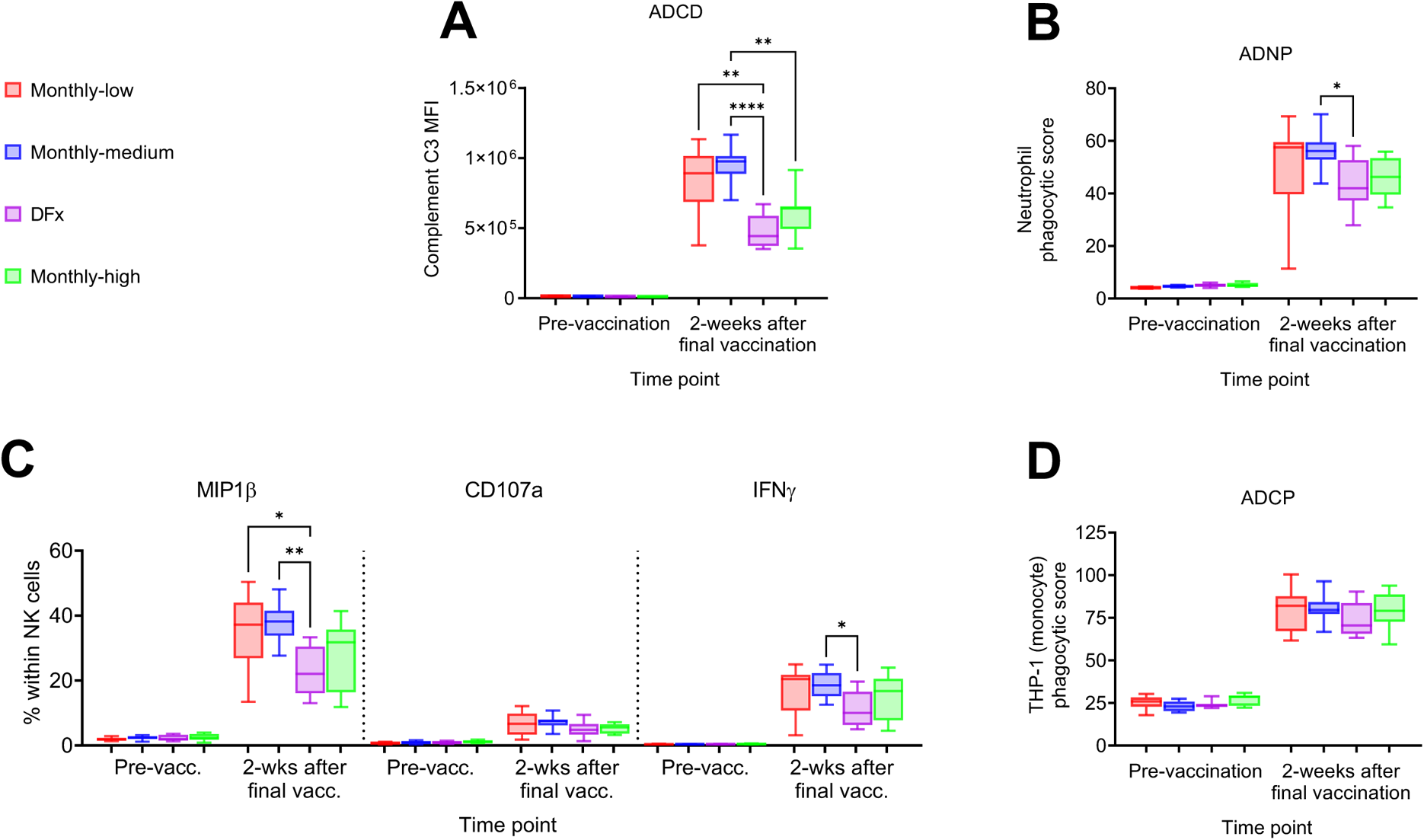
Fc-mediated functionality of peak post-vaccination antigen-specific Ig in DFx and monthly dosing regimens. Plasma from pre-vaccination and 2-weeks following the final vaccination was assessed for the capacity of anti-PfRH5 Ig to induce Fc-mediated innate immune activation following incubation with PfRH5-coupled beads. The Fc-mediated functionality of post-vaccination anti-PfRH5 Ig was compared between dosing regimen with respect to (**A**) antibody-dependent complement deposition (ADCD), (**B**) antibody-dependent neutrophil phagocytosis (ADNP), (**C**) NK cell activation, and (**D**) antibody-dependent cellular (monocyte) phagocytosis (ADCP). (**A**) Beads were incubated with guinea pig complement and C3 complement deposition was detected by staining with an anti-C3 fluorescent antibody and reported with the median anti-C3 fluorescence intensity (MFI) of each sample. (**B**) Neutrophils were isolated from fresh blood and incubated with beads, then stained to define neutrophils as SSC^hi^CD66b+CD14-CD3-cells. Induction of phagocytosis was compared by calculating phagocytic scores as (% bead-positive cells) x (gMFI of bead-positive cells) / (10 x gMFI of the first bead-positive peak). (**C**) NK cells were purified from buffy coats and incubated with antigen-coated ELISA plates, then stained to define NK cells as CD56+CD3-cells. Activation was measured as the percentage of NK cells expressing MIP1β, CD107a, or IFN-γ as detected by fluorescent antibodies. (**D**) The functional capacity of pre-/post-vaccination plasma to induce antibody-dependent monocyte phagocytosis was compared based on the capacity of anti-PfRH5-bound beads to induce phagocytosis by the THP-1 (monocyte) cell line. Phagocytic score of each sample = (% bead-positive cells) x (geometric median fluorescence intensity [MFI]) / (10x MFI of first bead-positive peak). Plasma was available from all vaccinees for inclusion in these analyses in technical duplicates. Pre-vaccination/ post-vaccination: Monthly-low *n* = 12/12; monthly-medium *n* =11/11; DFx *n* = 11/11; monthly-high *n* = 9/9. Comparisons between groups were performed by Kruskal-Wallis test with Dunn’s correction for multiple comparisons. Whiskers denote 5^th^ and 95^th^ percentiles. * *p* < 0.05, ** *p* < 0.01, **** *p* < 0.0001.

To deconvolute these data, we took three approaches with our subsequent computational analyses of the complete systems serology datasets. First, we focused on our original research question by comparing all monthly versus DFx regimens (**Figure 3A-C**). Next, we limited this analysis to a direct comparison between the DFx and monthly-high vaccinees, these differ only in their final vaccination and are thus optimal comparators (DFx versus monthly-high; **Figure 3D-F**). Finally, we addressed the new hypothesis raised by the univariate Fc functional data and compared the monthly-low and monthly-medium regimens to the monthly-high and DFx regimens (**Figure 3G-I**). The computational pipeline in each instance consisted of performing a partial least-squares discriminant analysis (PLS-DA) using features selected via least absolute shrinkage and selection operator (LASSO; **Figure 3A-B, D-E, G-H**), and finally building Spearman correlation networks to reveal additional serology features significantly associated with the selected features (**Figure 3C, F, I**). Parallel computational analyses correcting for total anti-RH5 IgG yielded comparable results as expected (data not shown), given there were no significant differences between regimens at the 2-week post-vaccination time point (16).

**Figure 3.**
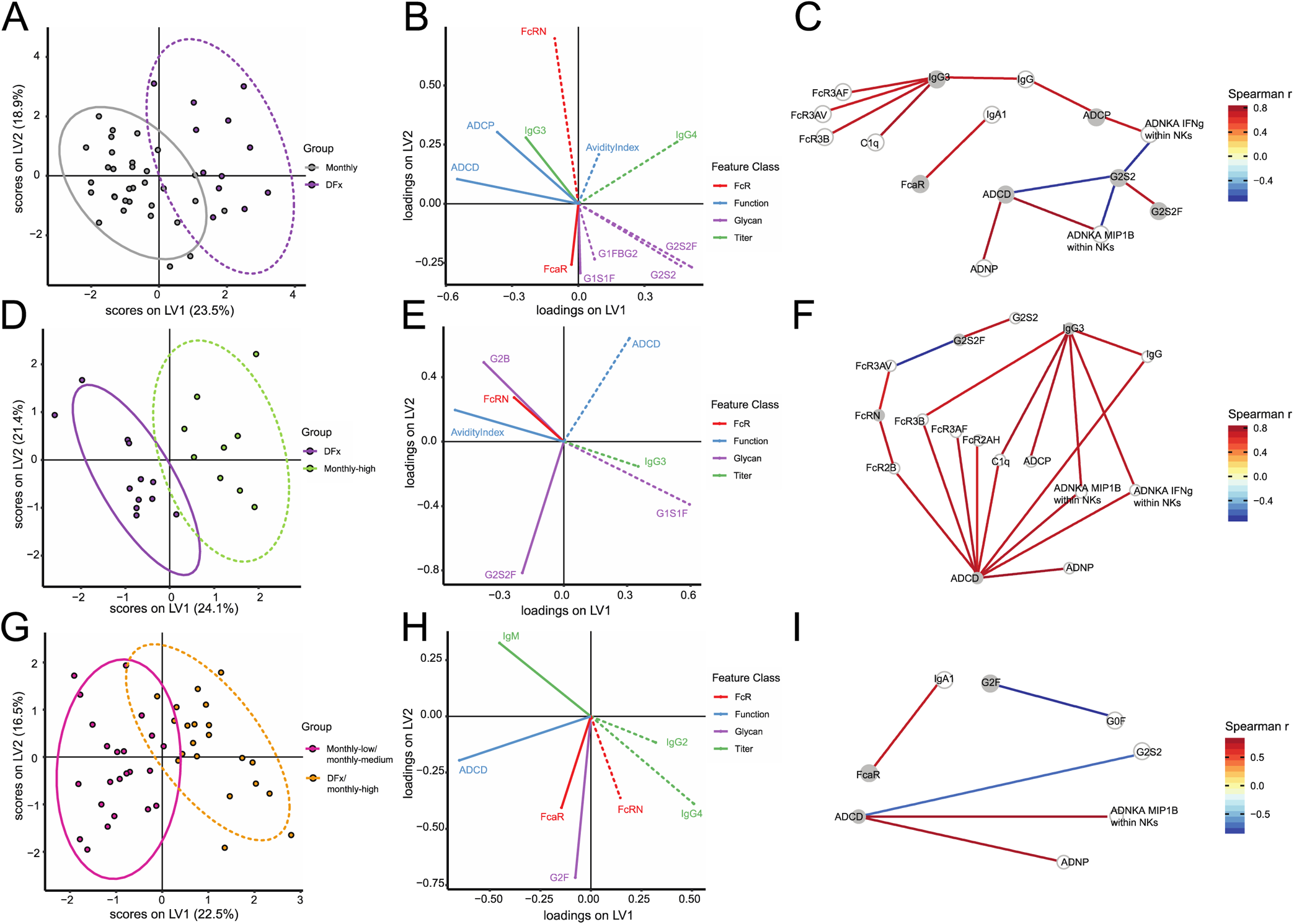
Systems serology computational analyses to define Ig feature sets that distinguish DFx from monthly dosing regimen. Partial least-squares discriminant analysis (PLS-DA) was performed with univariate read-outs from the systems serology analyses to compare DFx and monthly dosing regimens. Significant features were chosen via the LASSO feature selection algorithm and those chosen in at least 80% of 100 repetitions were used to build PLS-DA classifiers (**A-B**). Correlation networks were built to reveal additional serology features significantly associated with the selected features. Serology features significantly (p < 0.05, after a Benjamini-Hochberg correction) correlated via Spearman correlation (*r_S_* >|0.7|) were selected as co-correlates (**C**). The gradient colour of edges represents correlation value between the features, represented as nodes. Nodes are coloured according to selected status, with grey nodes as selected features and white nodes as co-correlate features. This approach was also used to directly compare DFx vaccinees with monthly-high (**D, E, F**) and DFx/ monthly-high vaccinees with monthly-low/ monthly-medium regimens (**G, H, I**). Line style (solid vs hash) of features in (**B, E, H**) relates to group with significant increase in that feature with 95% confidence interval. Models had cross-validation accuracies of 0.85 (**A**), 0.74 (**D**), and 0.83 (**G**), with comparisons to null models generated by random feature selection (*p* = 0.01, *p* = 0.09, *p* = 0.01 respectively) or permuted labels (*p* < 0.01, *p* = 0.04, *p* < 0.01 respectively) being significant for all but comparison to a null model built with randomly selected features in DFx vs monthly-high vaccinees. This is likely due to the limited number of samples and the high correlations between selected features and non-selected features (**F**). LV: latent variable; ADCD: antibody-dependent complement deposition; ADCP: antibody-dependent cellular (monocyte THP-1) phagocytosis; ADNP: antibody-dependent neutrophil phagocytosis; ADNKA: antibody-dependent NK cell activation. Monthly-low: *n* = 12. Monthly-medium: *n* = 11. DFx: *n* = 12, Monthly-high: *n* = 9.

The first analysis indicates a feature set able to discriminate between DFx and monthly regimens: FcRn-binding, IgG avidity, proportion of three different Fc glycans (G1FBG2, G2S2, G2S2F), IgG4 (all higher in DFx vaccinees); and IgG3, FcαR-binding, ADCD, and ADCP, and G1S1F glycan (all higher in monthly regimen vaccinees; **Figure 3B; Supplemental Figure 3**). The co-correlate network additionally detects a significant negative association between G2S2 and both NK cell (MIP1β and IFNγ) and complement cascade activation (**Figure 3C**). Conversely, ADCD and ADCP are associated with the monthly regimens, as suggested by the univariate data (**Figure 2**).

With respect to the second analysis, direct comparison of the DFx regimen with only the monthly-high regimen showed that increased FcRn-binding, IgG avidity, G2S2F and G2B glycans, alongside decreased IgG3, ADCD, and G2S1F glycan, were able to significantly discriminate DFx vaccinees (**Figure 3E; Supplemental Figure 3**). The corresponding co-correlate network also identified positive correlations between FcRn-binding and FcR2B− or FcR3AV-binding, as well as correlations between G2S2F and G2S2 (positive) and FcR3AV-binding (negative).

Finally, the computational analyses identified increased FcRn-binding, IgG2 and IgG4, in addition to decreased IgM, ADCD, FcαR-binding, and G2F glycan as the significant feature set to discriminate the ‘high’ dose vaccinees (**Figure 3H**). To note, the FcRn-binding signature in this analysis is attributable only to the DFx samples: FcRn-binding levels separate DFx vs monthly-high (**Figure 3E**) and removing FcRn-binding from the feature set, considered when separating lower vs higher dose groups, does not impact separation (data not shown). The selected features truly elevated in ‘high’ dosing are thus IgG2 and IgG4. This is consistent with the divergent IgG subclass immunokinetics analysed by ELISA described above (**Figure 1D-G**). No further correlations were identified in the co-correlates model with any of the three parameters elevated in the ‘high’ dose groups (**Figure 3I**).

### Single cell RNA sequencing indicates higher proportion of plasma cells in monthly boosting regimen antigen-specific B cell population 2-weeks following final vaccination as compared to DFx dosing regimen

Taken all together, and alongside previously published complementary observations on serum anti-PfRH5 IgG avidity and longevity (1), the above data indicate substantial quantitative and qualitative differences between the PfRH5-specific humoral responses following monthly or DFx regimens. These results infer disparate B cell activation following the final vaccination, with downstream impact on the phenotype and longevity of antibody-secreting B cells.

While we had already shown that the DFx regimen induces a higher frequency of circulating CD19+IgG+ PfRH5-specific B cells (memory cells as shown in **Figure 1B**), it was not clear whether there were any qualitative differences within this population that might explain differences in the humoral response described above. To address this, we performed scRNA-seq with single cell sorted CD19+IgG+ PfRH5-specific B cells from *n*=4 monthly-high vaccinees and *n*=3 DFx vaccinees using a method based on Smart-Seq v4 for Ultra Low Input RNA. The post-vaccination time point with the highest frequency of PfRH5-specific cells (2-weeks after final vaccination; **Figure 1B**) was selected to maximise the number of cells available for sequencing. The range of frequencies of PfRH5-specific B cells within the live, CD19+IgG+ population acquired during sorting (**Figure 4A**; monthly regimen 0.6-1.9%, DFx regimen 1.7-8.9%) were comparable to those previously observed (**Figure 1B**).

**Figure 4.**
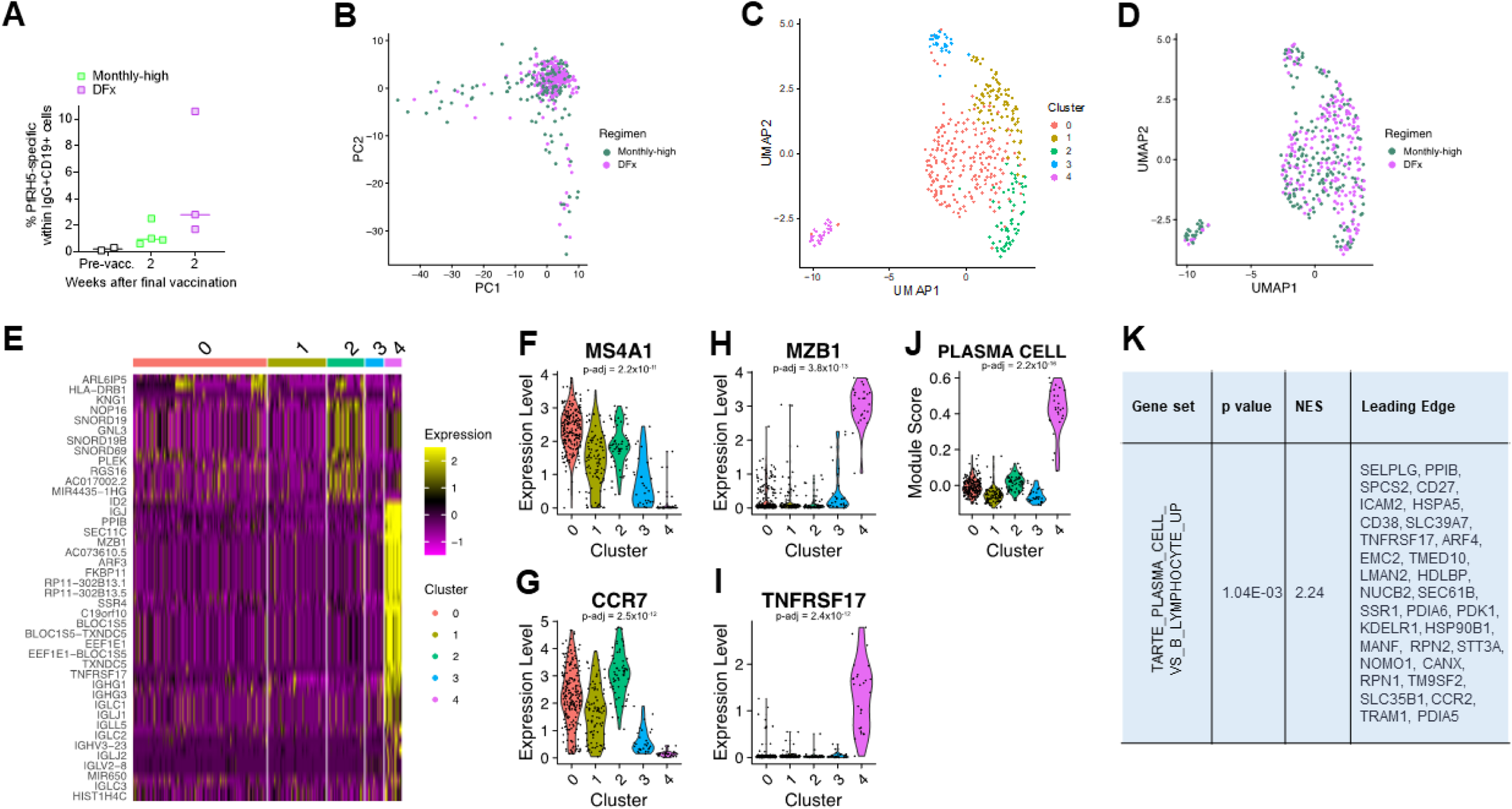
Single cell RNA sequencing of antigen-specific B cells from DFx and monthly-high dosing vaccinees. PBMC from pre-vaccination (Pre-vacc.), and 2-weeks post final vaccination in DFx vaccinees (*n*=3) and monthly-high vaccinees (*n*=4) were enriched for B cells and then stained with phenotypic markers for single cell sorting of antigen-specific B cells as defined as: live CD19+IgG+ lymphocytes that co-stained for monobiotinylated-PfRH5-PE and monobiotinylated-PfRH5-APC (gating strategy shown in **Supplemental Figure 1A**). Frequencies of post-vaccination PfRH5-specific B cells from the samples sorted were comparable with previous data (**A**). Libraries were sequenced following a Smart-Seq v4 and Nextera XT pipeline on a HiSeq4000. Variation in gene expression within the 7 samples was explored in Seurat by (**B**) PCA analysis of DFx as compared to monthly-high regimen, (**C**) UMAP with 5 clusters, (**D**) UMAP with 5 clusters with dosing regimen identity overlaid, and (**E**) heat map of the top 50 most differentially expressed genes by cluster. Expression of genes of interest identified were then compared between cluster 4 and the other clusters by Wilcoxon Rank Sum Test with Bonferroni correction to give adjusted *p* values (padj): (**F**) MS4A1 (CD20), (**G**) CCR7, (**H**) MZB1, and (**I**) TNFRSF17. Expression of a plasma cell gene set (“TARTE_PLASMA_CELLS_VS_B_LYMPHOCYTE_UP” (32)) was also compared between cluster 4 and the other clusters by Kruskal-Wallis test (**J**). Finally, gene set enrichment analysis with the TARTE_PLASMA_CELLS_VS_B_LYMPHOCYTE_UP gene set was run with the 5,115 significant genes from DESeq2 analyses comparing gene expression in PfRH5-specific CD19+IgG+ B cells from monthly-high and DFx regimen vaccinees (top twenty genes shown in Table 2). NES: normalised enrichment score; Leading Edge: subset of genes from gene set contributing most to the enrichment signal.

**Table 2.**
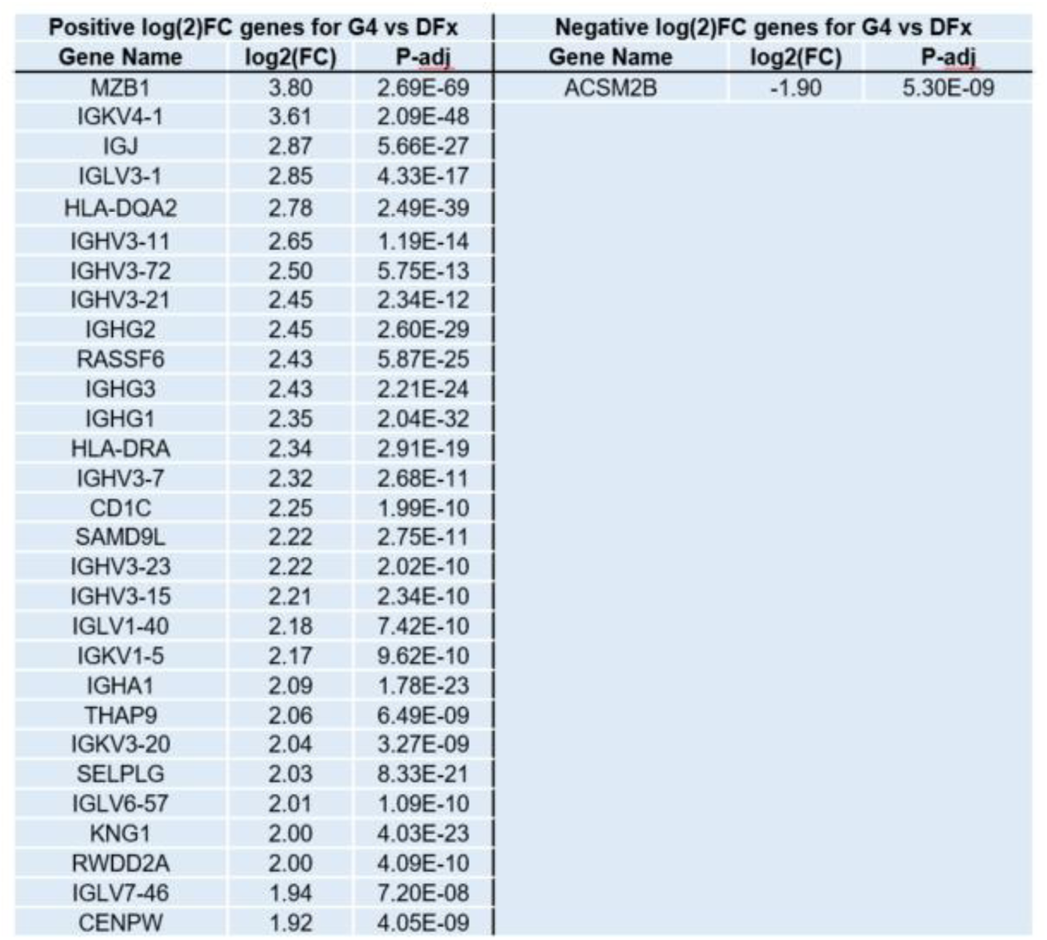
Differentially expressed antigen-specific B cell genes between DFx and monthly-high dosing vaccinees. Differentially expressed genes between DFx and monthly dosing samples were identified by DESeq2 and filtered to remove pseudogenes, LincRNA, IncRNA, and non-significant genes as defined as padj > 0.05. Genes were then ranked by fold change to show the top thirty genes differentially expressed between monthly-high regimen and DFx. Log2(FC): log fold change between monthly-high regimen and DFx; padj: adjusted *p* value after Benjamini-Hochberg correction for multiplicity of testing.

| Positive log<sub>2</sub>(FC) genes for G4 vs DFx |  |  | Negative log<sub>2</sub>(FC) genes for G4 vs DFx |  |  |
| --- | --- | --- | --- | --- | --- |
| Gene Name | log<sub>2</sub>(FC) | P-adj | Gene Name | log<sub>2</sub>(FC) | P-adj |
| MZB1 | 3.80 | 2.69E-69 | ACSM2B | -1.90 | 5.30E-09 |
| IGKV4-1 | 3.61 | 2.09E-48 |  |  |  |
| IGJ | 2.87 | 5.66E-27 |  |  |  |
| IGLV3-1 | 2.85 | 4.33E-17 |  |  |  |
| HLA-DQA2 | 2.78 | 2.49E-39 |  |  |  |
| IGHV3-11 | 2.65 | 1.19E-14 |  |  |  |
| IGHV3-72 | 2.50 | 5.75E-13 |  |  |  |
| IGHV3-21 | 2.45 | 2.34E-12 |  |  |  |
| IGHG2 | 2.45 | 2.60E-29 |  |  |  |
| RASSF6 | 2.43 | 5.87E-25 |  |  |  |
| IGHG3 | 2.43 | 2.21E-24 |  |  |  |
| IGHG1 | 2.35 | 2.04E-32 |  |  |  |
| HLA-DRA | 2.34 | 2.91E-19 |  |  |  |
| IGHV3-7 | 2.32 | 2.68E-11 |  |  |  |
| CD1C | 2.25 | 1.99E-10 |  |  |  |
| SAMD9L | 2.22 | 2.75E-11 |  |  |  |
| IGHV3-23 | 2.22 | 2.02E-10 |  |  |  |
| IGHV3-15 | 2.21 | 2.34E-10 |  |  |  |
| IGLV1-40 | 2.18 | 7.42E-10 |  |  |  |
| IGKV1-5 | 2.17 | 9.62E-10 |  |  |  |
| IGHA1 | 2.09 | 1.78E-23 |  |  |  |
| THAP9 | 2.06 | 6.49E-09 |  |  |  |
| IGKV3-20 | 2.04 | 3.27E-09 |  |  |  |
| SELPLG | 2.03 | 8.33E-21 |  |  |  |
| IGLV6-57 | 2.01 | 1.09E-10 |  |  |  |
| KNG1 | 2.00 | 4.03E-23 |  |  |  |
| RWDD2A | 2.00 | 4.09E-10 |  |  |  |
| IGLV7-46 | 1.94 | 7.20E-08 |  |  |  |
| CENPW | 1.92 | 4.05E-09 |  |  |  |

We first explored the heterogeneity in gene expression across all vaccinees within Seurat (**Figure 4B-J**). A total of 209 cells from monthly-high vaccinees and 214 cells from DFx vaccinees were included in this analysis. Following normalisation of data, we identified the ten genes with the highest variance across the entire sample set: MZB1, KNG1, LEO1, EAF2, UBXN8, P2RY12, ZNF234, MRPL35, TNFRSF17, IGKV1-39. Two of these genes are noteworthy for their associations with plasma cells that could be highly relevant for understanding our humoral data set: MZB1 and TNFRSF17. MZB1 – also known as Plasma cell-induced ER protein (pERp1) – is a key effector of the transcription factor B that regulates terminal plasma cell differentiation (Blimp1) and is of particular interest due to its reported roles in antibody secretion, as well as plasma cell differentiation and migration to the bone marrow (17–24). TNFRSF17 – also known as BCMA – is the APRIL and BAFF receptor which is restricted to mature B cells and plasma cells (including both short-lived and long-lived) and required for survival of LLPCs in the bone marrow (25–27).

Next, we ran a PCA to identify the number of principal components appropriate for downstream clustering and visualisation with UMAP (**Figure 4B**). Running UMAP on these principle components, we observe phenotypically distinct populations of cells (**Figure 4C**) one of which appeared to be enriched for monthly-high vaccinee cells (cluster 4; **Figure 4C-D**). The top 5 significant genes associated with this cluster (based on fold change) are IGHG2 (Ig Heavy Constant Gamma 2; i.e. heavy chain of IgG2), RP11-731F5.2 (LincRNA), IGLC1 (Ig Lambda Constant Region 1), IGLJ1 (Ig Lambda Joining 1), and IGLL5 (Ig Lambda Like Polypeptide 5; **Supplemental Table 1**). The other clusters are comprised more evenly of monthly-high regimen and DFx cells with the top genes for each as follows: MS4A1 (CD20) and YPEL5 for Cluster 0; GAREML and PSD for Cluster 1; LDHA and CCR7 for Cluster 2; and, MT-RNR2 and MUC3A for Cluster 3 (top 5 genes for each shown in **Supplemental Table 1**). While these gene rankings don’t suggest specific B cell subsets for clusters 0-3, the prevalence of Ig genes in cluster 4 suggest this may be an (antibody-producing) plasma cell population. A heat map visualisation of the top 50 most differentially expressed genes between the clusters additionally highlights increased expression among nearly all cells in cluster 4 of further genes with potential relevance to Ig production or plasma cell phenotype. In addition to MZB1 and TNFRSF17, this group of genes includes multiple encoding proteins with functions related to the secretory pathway or protein production and trafficking e.g. PPIB, ARF3, FKBP11, SEC11C, SSR4, BLOC1S5, and TXNDC5 (**Figure 4E**).

To further probe this potential cluster 4 plasma cell phenotype, we plotted several of the genes of interest as flagged above that should have clear positive or negative expression in a plasma cell population: MS4A1 (CD20), CCR7, MZB1 and TNFRSF17. CD20 and CCR7 are both downregulated during plasma cell differentiation (20, 28–30) and indeed here we see decreased expression in cluster 4 as compared to clusters 0-3 (**Figure 4F-G**). Given that the absence of CD20 expression on a B cell is the canonical definition of an antibody-secreting cell (though other marker combinations can be used e.g. CD27hiCD38hi (28)), this is strong evidence that cluster 4 is a plasma cell population. CD20 expression has also been used previously to differentiate antibody-secreting (plasma) cells from other probe-specific B cells – defined as activated B cells (ABCs) – following influenza vaccination (31). Conversely, both MZB1 and TNFRSF17 – both with clear plasma cell biological roles – are expressed almost exclusively in cluster 4 (**Figure 4H-I**). Finally, scoring each of the clusters based on expression of a previously identified set of genes upregulated in plasma cells as compared to other B cells (“TARTE_PLASMA_CELLS_VS_B_LYMPHOCYTE_UP” (32)) also yields a strong statistically significant difference between cluster 4 and the other clusters (**Figure 4J**).

We next proceeded to validate the indication from this single cell Seurat analysis that there was a discrepancy in the proportion of plasma cells (cluster 4) within PfRH5-specific B cells by dosing regimen using differential gene expression analyses between monthly-high and DFx regimen vaccinees. A total of 5,115 genes were differentially expressed (adjusted p value (padj) <0.05); the 30 significant genes with the greatest fold change in expression are presented in **Table 2**. Genes with increased expression in cells from monthly-high as compared to DFx vaccinees include MZB1, IGJ (also known as J Chain and associated with plasma cells (33), HLA-DQA2, and HLA-DRA. Of the other 25 genes, 17 were immunoglobulin heavy or light chain likely indicating increased Ig production. To note, while it is also possible that the Ig genes merely reflect differences between vaccinees in Ig gene usage, a similar increase in different Ig heavy or light chains is not observed in the DFx vaccinees. Only one gene downregulated in monthly-high vaccinees as compared to DFx is included in the top 30 differentially expressed genes: ACSM2B, which is involved in first step of fatty acid metabolism. As germinal centre B cells primarily utilise fatty acid metabolism to meet their metabolic needs – unlike other proliferating B cells (34, 35) – this could be indicative of a difference in germinal centre kinetics and output between the two regimens.

Further exploration of the significantly differentially expressed genes by gene set enrichment analysis (GSEA) with the Kegg mSigDB gene sets indicated significant enrichment (padj<0.05) in 10 pathways, including: cell adhesion molecules, protein export, and intestinal immune network for IgA production (**Supplemental Table 2**). Protein secretion and mTORC1 signalling (also related to protein secretion) were similarly flagged during Hallmark GSEA but no pathway reached statistical significance with an adjusted p value (**Supplemental Table 3**). A third GSEA with the plasma cell gene set used in **Figure 4J** – here run on all significantly differentially expressed genes rather than just cluster 4 – also showed a significant enrichment (p<0.0001; NES=2.24; **Figure 4K**).

Initially, the remaining disease-related pathways (**Supplemental Table 2**) do not seem to be of clear relevance to vaccine-specific B cell responses. However, the leading edges of each gene set (i.e. the subset of genes contributing most to the enrichment signal) reveal a central role for HLA class II genes in all pathways not otherwise linked to protein – putatively, immunoglobulin – production. While data on HLA-DQ and HLA-DP genes is scarce, HLA-DR is often used as a marker of activation, particularly of proliferating plasmablasts (i.e. SLPCs) as opposed to more mature LLPCs or other mBC phenotypes (28, 36, 37). The dominance of HLA class II genes in the leading edges of pathways enriched in monthly regimen vaccinees is therefore consistent with a higher proportion of SLPCs within the circulating PfRH5-specfic B cell pool 2-weeks following final vaccination as suggested by the Seurat analyses (**Figure 4**) and differentially expressed genes highlighted in the original DESeq2 analyses (**Table 2**).

### PfRH5-specific B cells show greater CDR3 somatic hypermutation in DFx vaccinees as compared to monthly regimen vaccinees

As the polyclonal serum anti-PfRH5 IgG antibody analyses demonstrated higher avidity in the DFx vaccinees (1), we were also interested to interrogate the BCR repertoire of the circulating PfRH5-specific B cells to determine if there were differences in percentage germline mutation or other divergent trends in clonotypes. Following analysis with the MiXCR pipeline to extract CDR3 sequences from the scRNA-seq data set, we compared CDR3 length as well as heavy and light chain CDR3 V-(D)-J percentage mutation from germline between monthly-high and DFx vaccinees. Here, we observe minimal differences in CDR3 length (**Figure 5A**), but increased somatic hypermutation in the DFx regimen PfRH5-specific cells as compared to those from the monthly-high regimen (**Figure 5B-C**). Analysis of heavy chain V gene usage showed substantial variation between individuals (**Supplemental Figure 4**) but broadly similar patterns when comparing monthly-high regimen and DFx vaccinee groups (**Figure 5D-E**). A total of 40 V genes were detected in CDR3 heavy chains, with 5 of the 6 top genes the same in both monthly-high and DFx vaccinees (IGHV4-39, IGHV4-31, IGHV3-23, IGHV3-21, and IGHV3-33). Finally, hierarchical clustering was performed with heavy chain CDR3 amino acid sequences using Geneious Tree Builder and visualised as unrooted dendrograms (**Figure 5F-G; Supplemental Figure 4**). Three clusters were observed in both monthly-high regimen (**Figure 5F**) and DFx vaccinees (**Figure 5G**), as well as individual vaccinees (**Supplemental Figure 4**).

**Figure 5.**
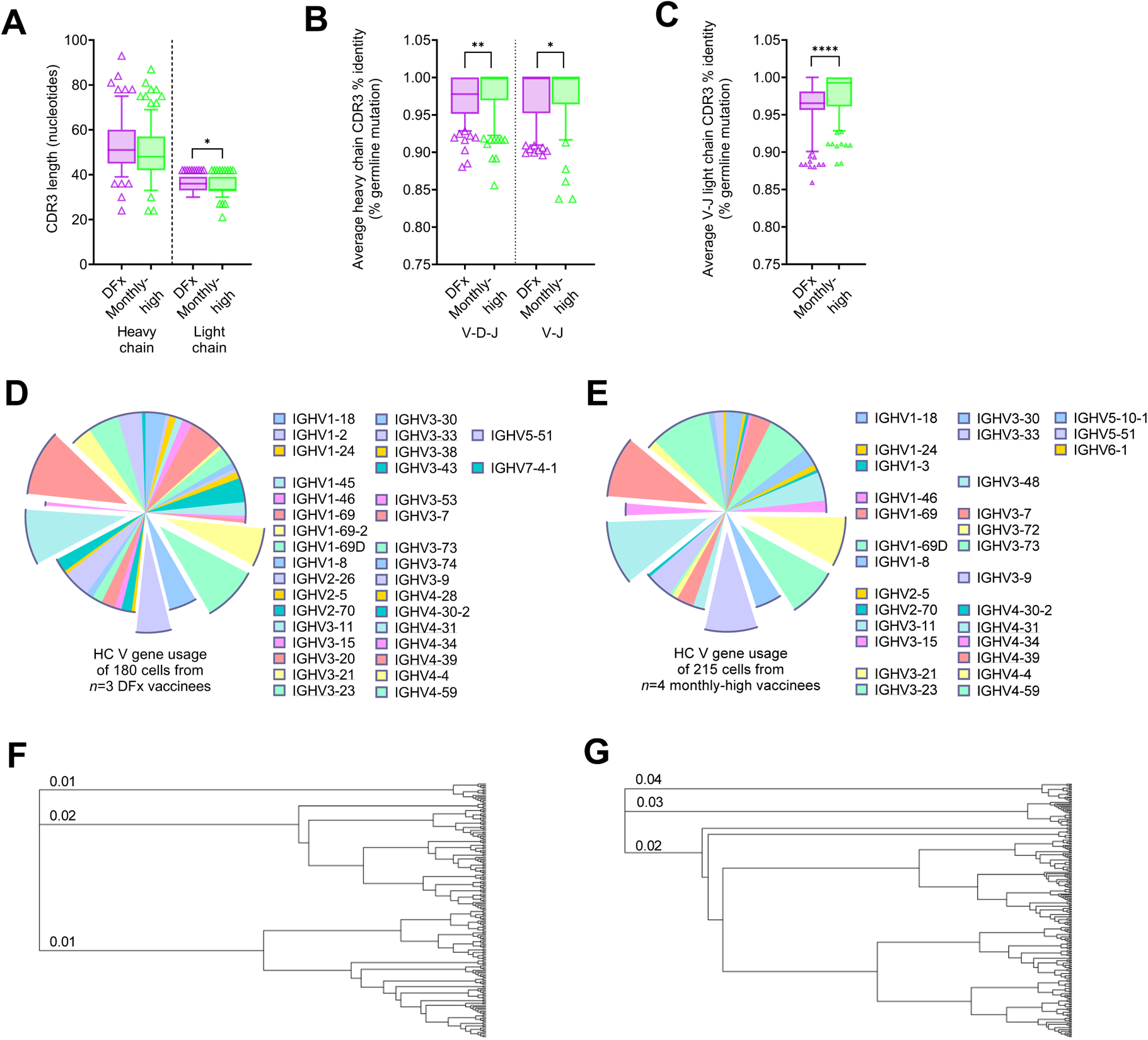
CDR3 sequence analysis of PfRH5-specific B cells in DFx and monthly-high dosing vaccinees. PBMC from pre-vaccination (Pre-vacc.), and 2-weeks post final vaccination in DFx vaccinees (*n*=3) and monthly-high vaccinees (*n*=4) were enriched for B cells and then stained with phenotypic markers for single cell sorting of antigen-specific B cells as defined as: live CD19+IgG+ lymphocytes that co-stained for monobiotinylated-PfRH5-PE and monobiotinylated-PfRH5-APC (gating strategy shown in **Supplemental Figure 1A**). Libraries were sequenced following a Smart-Seq v4 and Nextera XT pipeline on a HiSeq4000. CDR3 sequences were extracted using the MiXCR pipeline to compare heavy and light chain CDR3 lengths (**A**), and average percentage germline identity for V-D-J or V-J heavy chain (**B**) and V-J light chain (**C**) sequences. Comparisons were performed by Mann-Whitney tests. Whiskers denote 5^th^ and 95^th^ percentiles. * *p* < 0.05, ** *p* < 0.01, **** *p* < 0.0001. Heavy chain (HC) V gene usage is reported for monthly-high (**D**) and DFx (**E**) vaccinees. The top five HC V genes shared between groups (IGHV3-21, IGHV3-23, IGHV3-33 IGHV4-31, and IGHV4-39,) are emphasised in the charts and underlined in the key. Hierarchical clustering was performed on CDR3 HC amino acid sequences and visualised as dendrograms for monthly-high vaccinees (**F**), and DFx vaccinees (**G**). The first three branches are labelled in each dendrogram with branch length (distance between internal nodes) i.e. substitutions per amino acid.

## Limitations

As is often true for scRNA-seq analyses, our sample size is relatively small. Here, we compared gene expression and BCR repertoires between *n*=4 monthly regimen vaccinees and *n*=3 DFx vaccinees. Similarly, while the sample size for the systems serology work was higher (*n*=32 monthly regimen; *n*=12 DFx) our conclusions from these data sets could again be strengthened with future replication studies with further vaccinees. Reproducing our findings with similar dosing regimens but different antigens would be particularly of interest to confirm translatability of our findings.

It is also important to note two potential limitations associated with the use of the PfRH5 probes and the gating strategy used to define PfRH5-specific B cells for the scRNA-seq pipeline. First, while we routinely observe negligible background probe staining with Day 0 samples (**Figure 1**; **Supplemental Figure 1**) we did not clone monoclonal antibodies (mAbs) from BCRs of single sorted cells and screen to verify PfRH5 specificity. It is therefore possible that a minority of cells included in the sequencing analysis are not truly PfRH5-specific. Second, while previous work in our lab (data not shown) has demonstrated that CD19+IgG+ gating, during single cell sorting, facilitates clear differentiation between probe-negative and probe-positive populations, it is important to note that any downstream analyses consequently exclude any CD19-B cell populations as well as IgM+ or IgA+ subsets. Likewise, IgG+ plasma cell populations with very low BCR expression may have been missed from the IgG+ gate.

## Discussion

In this study we have interrogated the circulating antigen-specific Ig and B cell responses following immunisation with either monthly (0-1-2 month) or DFx (0-1-6 month) dosing regimens using the same vaccine. To the best of our knowledge, this is the first detailed exploration of B cell and humoral responses to a vaccine regimen that significantly improves Ig durability in humans. We observe that the DFx regimen greatly enhances the longevity of the IgG1 component of the total IgG response, with additional indications of improved serum maintenance across multiple other subclasses and isotypes (IgG3, IgA, IgA1, IgM). While the serum antibody kinetics are unique to DFx, analysis of peak Ig responses (2-weeks following final vaccination) suggested some similarities with the monthly regimen group with the same ‘high’ (50µg) first and second doses (monthly-high: 50-50-50µg). Specifically, these two groups of vaccinees shared increased IgG2 and IgG4 titres, in comparison to the monthly-low and monthly-medium groups. Univariate analyses of FcR-mediated functionality at the peak post-vaccination time point indicated other similarities between DFx and monthly-high regimen, with significantly lower activation of ADCD, and trends towards reduced ADCP and antibody-dependent NK cell activation as compared to monthly-low / monthly-medium. Systems serology computational analyses indeed identified IgG2 and IgG4 as components of the feature set that was associated with ‘high’ DFx / monthly-high.

An increase in antigen-specific IgG4 following DFx has been observed previously with the PfCSP-based pre-erythrocytic malaria vaccine RTS,S (0-1-7mo in this instance; Fx017M) when compared to a monthly 0-1-2mo regimen, as well as an IgG4-related decrease in ADCP (38). In the context of PfCSP-mediated vaccines it appears that IgG4 inhibition of opsonophagocytosis has a positive impact on protection, potentially related to a sporozoite immune escape mechanism, but whether this would also be true for blood-stage parasites is unknown. The authors comment that the prevailing view of chronic exposure to antigen as a driver of IgG4 (reviewed in (39, 40)) seems to conflict with their data, whereas the relevance appears clearer in our PfRH5 analyses whereby IgG4 is increased in both the DFx and monthly-high groups primed with the highest 50 µg dose of RH5.1 vaccine. To the best of our knowledge, there are no published data on the impact of relatively ‘high’ protein vaccine dosing on induction of an IgG2 response and the interpretation of these results is not clear.

With respect to the DFx regimen specifically, the associated systems serology feature set, as compared to all monthly regimen groups, consisted of increased FcRn-binding, IgG avidity, IgG4, and proportions of three galactosylated Fc glycans (G1FBG2, G2S2, G2S2F) and decreased IgG3, ADCD, ADCP, FcαR-binding, and G1S1F galactosylated Fc glycan. This was refined when analyses were limited to a DFx versus monthly-high regimen comparison: elevated FcRn-binding, IgG avidity, G2S2F and G2B Fc glycans, and decreased ADCD, IgG3, and G1S1F glycan. Our data therefore suggest both a negative impact of ‘high’ first and second doses (e.g. reduced capacity to induce ADCD) and a positive impact of the delayed fractional final dose (e.g. increased avidity and FcRn-binding) on the antigen-specific Ig response with the DFx regimen. This concept of a potential detrimental impact of excessive antigen or antigen saturation on vaccine immunogenicity has previously been proposed by others and merits further investigation, as do DFx regimens using lower priming doses (9, 41).

The avidity signal for DFx bolsters the conclusions of our previously published trial report with a different assay (1), while the glycan, subclass and FcRn-binding data provide novel insights into the profound modulation of the humoral response that can be achieved through modification of dosing regimen. The importance of improved avidity in the specific context of PfRH5-based vaccines is uncertain given previous work with mAbs (derived from samples from the viral vector trial) indicated that the speed of antibody-binding, rather than avidity, is more relevant for *in vitro* anti-parasitic functionality (1, 42). However, improved antibody avidity is associated with increased protection against other pathogens (including pre-erythrocytic *P. falciparum* malaria (7, 43–46)) and thus these DFx data may be of great interest to other vaccine development programmes. Importantly, this serum avidity improvement is supported by CDR3 sequence analysis of single cell sorted PfRH5-specific B cells where greater somatic hypermutation was observed in DFx vaccinees as compared to monthly regimen vaccinees. Further studies are also underway to interrogate the binding kinetics of mAbs derived from monthly regimen and DFx RH5.1/AS01_B_ vaccinees and the relationship to the *in vitro* functional correlate of protection (growth inhibitory activity [GIA] (1)).

The biological implication of increased capacity of post-vaccination Ig to bind to FcRn is noteworthy due to the central role of FcRn-binding in antibody longevity through promoting recycling rather than lysosomal degradation of IgG in circulation; durability of mAbs can be enhanced by modifying the Fc to improve FcRn-binding (47, 48). FcRn-binding can also be ameliorated with more highly galactosylated Fc glycans (15) and, consistent with this, the systems serology signature associated with the DFx samples included increased proportions of two bi-galactosylated glycan moieties (<u>G2</u>B and <u>G2</u>S2F). Similarly, and consistent with the avidity data, is evidence from influenza vaccine antibody analyses demonstrating increases in sialylation (e.g. as in G2<u>S2</u>F) correlated with generation of higher affinity antibodies (15, 49).

It is therefore possible that in DFx vaccinees there is an increase in expression of the glycosyltransferases responsible for adding galactose (B4GALT1) and sialic acid (ST6GAL1) with functional significance for FcRn-binding / longevity (galactose) and avidity (sialic acid). These hypotheses require further interrogation however, especially given the glycosylation changes are more specific than increases in all bi-galactosylated or bi-sialylated moieties. To note, although adjuvant selection can demonstrably impact peak antibody responses and antibody quality (including Fc glycans), evidence of any effect on antibody long-term maintenance is thus far limited to non-human primates (50, 51).

These changes in antigen-specific Ig quantity and quality thus suggest fundamentally different B cell responses following the third dose in DFx versus monthly regimens. Based on observed total IgG kinetics, we have previously hypothesised that the DFx regimen seeds a higher frequency of LLPCs in the bone marrow (1) but empirical data on direct detection of antigen-specific B cells (of any phenotype) in circulation or in lymphoid tissue have not been reported. We therefore first measured the frequency of peripheral antigen-specific cells within the memory IgG+ B cell population. Here, we showed that the frequency of circulating antigen-specific B cells was significantly higher in DFx vaccinees at all time points measured. However, we do not expect that a higher magnitude B cell response alone could be responsible for the range of qualitative differences in the humoral response detailed above. We therefore proceeded to scRNA-seq as an agnostic approach to qualitative antigen-specific B cell analysis with the goal of detecting any indication of discrepancies in B cell kinetics or fate decisions between the two dosing regimens. In light of the potential confounding factor of ‘high’ vs ‘low’ first and second doses, we restricted this analysis to DFx and monthly-high regimen.

Here, several pieces of data pointed to an increased proportion of plasma cells within the antigen-specific B cell population in monthly-high vaccinees, as compared to DFx. First, we identified a subpopulation of antigen-specific B cells that was almost exclusively comprised of cells from monthly-high vaccinees. This cluster was recognised as a plasma cell population due to the high expression of plasma cell markers MZB1 and TNFRSF17 (BCMA), negligible expression of MS4A1 (CD20) and CCR7, and significant enrichment for a pre-defined plasma cell gene set. Second, differential gene expression analysis (DESeq2) identified MZB1 as the gene with the highest fold change in expression between monthly-high and DFx samples, with 18 of the other top 29 genes comprised of IGJ or Ig heavy and light chains – consistent with increased antibody secretion. Indeed, in a recent single cell atlas characterisation of human tonsillar B cell populations, MZB1, IGJ and IGH genes were all expressed most highly in the plasmablast population (52). Finally, KEGG gene set enrichment analysis with the significantly differentially expressed genes highlighted the protein export pathway as significantly upregulated in monthly-high B cells as compared to DFx B cells.

At first glance, these scRNA-seq results suggesting greater IgG+ plasma cell activation in monthly-high than DFx are surprising, especially when anti-PfRH5 serum IgG levels are comparable between the two regimens at the peak time point used for this analysis (2-weeks post final vaccination (1)) and subsequently higher in DFx vaccinees long-term. However, selection of a single time point provides only a plasma cell “snapshot” rather than longitudinal kinetics; it is entirely feasible that the DFx response actually peaked between 2- and 4-weeks following final vaccination versus between 1- and 2-weeks with monthly-high. Furthermore, LLPCs exit later than mBCs in germinal centre development and, while this has been documented from 2-weeks post-vaccination onwards, it is possible that our scRNA-seq time point is better suited to mBC and SLPC detection as compared to (later) LLPCs (53–55). Accordingly, we propose that the Day 70 plasma cell signal in monthly-high vaccinees is derived predominantly from mBCs that have differentiated into short-lived plasma cells (SLPCs) following antigen re-exposure, rather than those that have returned to a draining lymph node and differentiated into LLPCs via germinal centre reactions. It is also likely that monthly regimen vaccinees still possessed PfRH5-specific germinal centres from the second vaccination at the time of the third and final booster which – alongside higher concentrations of anti-PfRH5 serum IgG at the time of vaccination – could have dampened new germinal centre responses (9, 56, 57). This is consistent with HIV vaccinology data from non-human primates where longer intervals between doses were associated with increased germinal centre B cell responses (58). Germinal centre-independent SLPC differentiation in monthly-high vaccinees would also be consistent with the observed lower CDR3 somatic hypermutation (and reduced serum IgG avidity) as compared to DFx vaccinees.

Another factor relevant for reconciling the serum IgG and plasma cell data is possible variation in CD19 expression by different plasma cell populations. CD19 is a common marker for identifying human B cells, but it has also been reported to be downregulated on terminally differentiated bone marrow LLPCs as well as on a minority of circulating plasma cells following vaccination (59–62). If peripheral LLPCs or LLPC-precursors downregulate CD19 they would not have been captured for the scRNA-seq pipeline by the CD19+IgG+ gating strategy if truly negative for CD19 (rather than CD19lo). This would again mean that the plasma cell population identified in our analyses is skewed to SLPCs, while any differences in germinal centre LLPC/precursor output between the two platforms are obscured. However, the significance or likelihood of circulating CD19-LLPCs and LLPC-precursors is unclear, given other reports of a putative LLPC-precursor population within the circulating CD19+ compartment (63) and timing of CD19 downregulation only once cells reached the bone marrow (59).

Future clinical trials should therefore seek to more precisely define this plasma cell population kinetics with DFx versus monthly boosting and to confirm our hypothesised skew towards LLPC and SLPC subsets, respectively. This distinction is currently difficult given the lack of clear markers to resolve SLPCs from LLPCs / LLPC precursors in humans (reviewed in (64)), although data suggest the transcription factor Zbtb20 may be used to define LLPCs in mice ((65) no increased expression was observed in our putative SLPC population). Moving forward will rely on more frequent venous sampling following the final vaccination, ideally coupled with analysis of draining lymph node aspirates to directly monitor germinal centre formation / longevity and define peripheral biomarkers of plasma cell output (57, 66, 67). Bone marrow aspirates may also be of use to confirm a link between higher serum anti-PfRH5 IgG maintenance and presumed higher LLPC seeding in the DFx regimen, as well as with putative LLPC precursor populations.

It will also be of great interest to better understand the discrepancy between the results observed with DFx RH5.1/AS01_B_ vaccination and similar Fx017M dosing with the RTS,S/AS01 which gave greater protection than the monthly regimen in controlled human malaria infection (CHMI) studies (6–8, 41). Like with DFx RH5.1 dosing, the Fx017M RTS,S regimen increased IgG avidity and (bulk plasmablast, i.e. SLPC) B cell somatic hypermutation after final vaccination – which correlated with protection – but not magnitude of the IgG response as compared to the monthly 0-1-2-month regimen (7). However, there was no apparent Fx017M-mediated benefit to Ig longevity and thus durability of vaccine-mediated protection (7). This is a critical distinction to understand in order to ensure relevance of DFx dosing to other antigens and pathogens. However, the failure of Fx017M to improve Ig serum maintenance could be related not to characteristics of the antigen, but of the platform; CSP is arrayed on a VLP which could potentially form immune complexes affecting LLPC development (68). Modelling data indeed indicate that IgG concentration and avidity are regulated independently by separate biological processes (69). Other discrepancies are also found with recently published systems serology analyses of monthly versus Fx017M dosing (70) where different trends were observed with Fx017M to those reported here with DFx.

In conclusion, the data presented here support the DFx regimen as a more promising dosing schedule as compared to more routine monthly dosing for optimising the humoral response against difficult pathogens like blood-stage malaria that require high, sustained titres for protection. The impact appears to be largely related to IgG1 but more subtle effects on other isotypes and subclasses are also present. Two hypotheses regarding the underlying mechanism of the improved plasma antibody longevity in the DFx schedule merit further exploration: increased recycling through enhanced FcRn-binding, and a potential shift in B cell fate from SLPC to LLPC following the delayed final dose. The mechanism for improved avidity with DFx is also likely linked to germinal centre kinetics, potentially related to competition for antigen which others have speculated is linked to the fractionation of the final dose rather than the delay (7, 41, 71, 72). Further clinical trials will therefore be needed to directly compare the impact of delayed boosting to the fractionation of the final dose, and also – for antigens other than PfRH5 such as PfCSP – to delineate the possible roles of antigen and vaccine delivery platform.

## Methods

### Resource availability

#### Lead contact

Further information and requests for resources and reagents should be directed to and will be fulfilled by the Lead Contact, Carolyn Nielsen.

#### Data and code availability

The scRNA-seq data generated during this study are available from the NCBI SRA database, BioProject accession number SUB10783850.

### Experimental model and subject details

This study focused on the comparison of immune responses between groups receiving different dosing regimens of PfRH5 protein (RH5.1) with AS01_B_ adjuvant (ClinicalTrials.gov Identifier: NCT029271452 (1, 12); adjuvant provided by GSK). The clinical trial NCT029271452 from which samples were used for this study was approved by the Oxford Research Ethics Committee A in the UK (REC reference 16/SC/0345) as well as by the UK Medicines and Healthcare products Regulatory Agency (MHRA; reference 21584/0362/001-0001). All volunteers gave written informed consent.

Vaccine regimens are presented in **Table 1**. Antibody and Tfh cell responses to the different regimens have already been reported elsewhere (1, 10). Vaccinee age and sex were comparable between regimens and are summarised in **Table 1**. Not all vaccinees could be run in each of the assays in this study; specific sample sizes are specified in figure legends.

A second PfRH5 clinical trial with heterologous viral vectors (consisting of a ChAd63-PfRH5 prime, followed by a MVA-PfRH5 boost) is also briefly referenced for comparison in **Figure 1** (ClinicalTrials.gov Identifier: NCT02181088; (10, 11). This trial was also approved by the Oxford Research Ethics Committee A in the UK (REC references 14/SC/0120) as well as by the UK MHRA. All volunteers gave written informed consent.

### Method details

#### PfRH5-specific B cell flow cytometry

Cryopreserved PBMC were thawed into R10 media (RPMI [R0883, Sigma] supplemented with 10% heat-inactivated FCS [60923, Biosera], 100U/ml penicillin / 0.1mg/mL streptomycin [P0781, Sigma], 2mM L-glutamine [G7513, Sigma]) then washed and rested in R10 for 1h. B cells were enriched (Human Pan-B cell Enrichment Kit [19554, StemCell]) and then stained with viability dye FVS780 (565388, BD Biosciences). Next, B cells were stained with anti-human CD19-PE-Cy7 (557835, BD Biosciences), anti-human IgG-BB515 (564581), anti-human IgM-BV510 (563113), anti-human CD27-BV711 (564893), anti-human CD21-BV421 (562966), as well as two fluorophore-conjugated PfRH5 probes. Preparation of the PfRH5 probes has been published previously (10). In brief, monobiotinylated PfRH5 was produced by transient co-transfection of HEK293F cells with a plasmid encoding BirA biotin ligase and a plasmid encoding a modified full-length PfRH5. The PfRH5 plasmid was based on ‘RH5-bio’ (gift from Gavin Wright [Addgene plasmid # 47780; http://n2t.net/addgene:47780;RRID:Addgene_47780] (73). RH5-bio was modified prior to transfection to incorporate a ‘C-tag’ for subsequent protein purification, as well as a 15 amino acid deletion at a predicted cleavage site. Probes were freshly prepared for each experiment, by incubation of monobiotinylated PfRH5 with streptavidin-PE (S866, Invitrogen) or streptavidin-APC (eBioscience, 405207) at an approximately 4:1 molar ratio to facilitate tetramer generation and subsequently centrifuging to remove aggregates. Following surface staining, cells were fixed with CytoFix/CytoPerm (554714, BD Biosciences), washed, and stored at 4°C until acquisition.

Memory PfRH5-specific IgG+ B cells identified as live CD19+CD21+CD27+IgG+IgM− PfRH5/APC+PfRH5/PE+ lymphocytes unless otherwise specified (**Supplemental Figure 1**) and acquired on a Fortessa X20 flow cytometer with FACSDiva8.0 (both BD Biosciences). Samples were analysed using FlowJo (v10; Treestar). Samples were excluded from analysis if <100 cells in the parent population.

#### Standardised ELISAs

Standardised ELISAs were used to quantify serum RH5.1-specific IgG1, IgG2, IgG3, IgA, IgA1, IgA2 and IgM responses in vaccinees. Nunc MaxiSorp™ flat-bottom ELISA plates (44-2404-21, Invitrogen) were coated overnight with 5µg/mL of RH5.1 protein in PBS. Plates were washed with washing buffer composed of PBS containing 0.05% TWEEN® 20 (P1379, Sigma-Aldrich) and blocked with 100µL of Blocker™ Casein in PBS (37582, ThermoFisher Scientific). After removing blocking buffer, standard curve and internal controls were created in casein using a pool of high-titre volunteer plasma, specific for each isotype or subclass being tested, and 50µL of each dilution was added to the plate in duplicate. Test samples were diluted in casein to a minimum dilution of 1:50 and 50µL was added in triplicate. Plates were incubated for 2 hours at 37°C and washed in washing buffer. An alkaline phosphatase-conjugated secondary antibody from Southern Biotech was diluted at the manufacturer’s recommend minimum dilution for ELISA in casein. The antibody used was dependent on the isotype or subclass being assayed and were as follows: Mouse Anti-Human IgG1 Fc-AP (9054-04), Mouse Anti-Human IgG2 Fc-AP (9060-04), Mouse Anti-Human IgG3 Hinge-AP (9210-04), Mouse Anti-Human IgG4 Fc-AP (9200-04), Goat Anti-Human IgA-AP (2050-04), Mouse Anti-Human IgA1-AP (9130-04), Mouse Anti-Human IgA2-AP (9140-04), Goat Anti-Human IgM-AP (2020-04). 50µL of the secondary antibody dilution was added to each well of the plate and incubated for 1 h at 37°C. Plates were developed using PNPP alkaline phosphatase substrate (N2765, Sigma-Aldrich) for 1-4 h at 37°C. Optical density at 405 nm was measured using an ELx808 absorbance reader (BioTek) until the internal control reached an OD_405_ of 1. The reciprocal of the internal control dilution giving an OD_405_ of 1 was used to assign an AU value of the standard. Gen5 ELISA software v3.04 (BioTek) was used to convert the OD_405_ of test samples into AU values by interpolating from the linear range of the standard curve fitted to a four-parameter logistics model. Any samples with an OD_405_ below the linear range of the standard curve at the minimum dilution tested were assigned a minimum AU value according to the lower limit of quantification of the assay.

### Systems serology

The PfRH5 systems serology analyses were performed as previously reported (1). A total of 49 parameters were measured in the assays outlined below: THP-1 phagocytosis, neutrophil phagocytosis, NK cell activation (3 read-outs), complement deposition, antibody isotype and subclass (9 read-outs including total IgG avidity), FcR-binding (8 read-outs), C1q binding, and frequencies of glycan structures (13 read-outs). These assays were performed at Massachusetts General Hospital (MGH) using plasma samples from the VAC063 trial and were deemed not human research following review by the MGH Institutional Review Board (protocol 2012P002452). Additionally, human whole blood and buffy coats were collected at MGH from healthy donors who did not participate in the VAC063 trial. Use of these internal samples as sources of uninfected primary neutrophils and NK cells was deemed not human research by the MGH IRB (protocols 2010 P002121 and 2005 P001218).

Details of individual assays are described in the Supplemental Materials.

### Single cell RNA sequencing of PfRH5-specific CD19+IgG+ B cells

Similar to as described for the B cell flow cytometry assay, here B cells were enriched (Human Pan-B cell Enrichment Kit [19554, StemCell]) from cryopreserved PBMC samples from two weeks after the final vaccination in three DFx vaccinees (50-50---10µg) and four monthly-high vaccinees (50-50-50µg). These samples were then stained with anti-human CD19-PE-Cy7 (557835, BD Biosciences), anti-human IgG-BB515 (564581), FVS780 (565388, BD Biosciences), as well as two fluorophore-conjugated PfRH5 probes (see flow cytometry section for more details on probe production (10)). PfRH5-specific B cells identified as live CD19+IgG+PfRH5/APC+PfRH5/PE+ lymphocytes were single cell sorted on a BD FACS Aria (BD Biosciences) into Buffer TCL (1031576, Qiagen) with 1% 2-mercaptoethanol. Sorted cells were snap frozen on dry ice before storage at −80°C until processing.

mRNA from thawed single B cells was purified with RNAClean XP beads (A63987, Beckamn Coulter) and converted to cDNA using dT_30_VN and TSO oligonucleotides and SMARTScribe reverse transcriptase (639538, Clontech) with a modified Smart-Seq v4 for Ultra Low Input RNA protocol (Takara Bio). Both steps were done in the presence of a recombinant RNase inhibitor (2313B, Takara Bio). cDNA was then amplified with SeqAmp DNA Polymerase (638509, Clontech).

HighPrep PCR beads (AC-60050, MagBio) were used to purify cDNA prior to quantification with Qubit dsDNA HS Assay Kit (Q32854, Life Technologies) and cDNA normalisation. Sequencing libraries were created using the Nextera Index Kit v2 (FC-131-2001, Illumina) and the Nextera XT DNA Sample Preparation Kit (FC-131-1096, Illumina). Libraries were then purified with AMPure XP beads (A63881, Beckman Coulter) and quantified by qPCR with Library Quantification Kit-Illumina/ ABI Prism (KK4835, KAPA Biosystems). Cells yielding libraries >1nM by qPCR were normalised and pooled by vaccinee for sequencing on the HiSeq4000 platform (Illumina) to an average read count of 4,113,948 reads per cell. All cells were sequenced in a single run. Sequencing was performed on a total of 226 cells from monthly-high vaccinees (range 37-88 cells) and 228 cells from DFx vaccinees (range 59-88 cells).

### Quantification and Statistical Analyses

#### Computational analysis of systems serology data

Multivariate analysis of the systems serology data was performed in R with the following approach. Features with missing measurements for more than 50% of subjects were removed from this analysis. Missing values were then imputed using k-nearest neighbors (k = 3, R package ‘DMwR’ v.0.4.1), and all data were mean-centred and variance-scaled (“z-scored”). Univariate differences were assessed with Kruskal-Wallis tests, using the Benjamini-Hochberg multiple hypothesis correction. Significant differences were then assessed in a pairwise manner with Mann-Whitney U tests. Partial least-squares discriminant analysis (PLS-DA) was performed using the ‘ropls’ (v.1.22.0) and ‘systemsseRology’ (v1.0) packages of R for model building and cross-validation/visualization respectively. Significant features were chosen via the LASSO feature selection algorithm, which was run 100 times on the entire dataset using the function ‘select_lasso’ from the ‘systemsseRology’ R package (https://github.com/LoosC/ systemsseRology). Features chosen in at least 80% of repetitions were used to build PLS-DA classifiers. PLS-DA model performances were then assessed using a five-fold cross validation approach and reported cross-validation accuracy is the mean of 10 rounds of 5-fold cross validation, which includes 100 repeats of feature selection per fold per round. To assess the importance of selected features, negative control models were built both by permuting group labels and by selecting random, size-matched features in place of true selected features. 10 rounds of cross-validation with 5 permutation and 5 random-feature trials per round were performed (again with 100 repeats of feature selection per fold per round for the permutation trials), and exact *p* values were obtained from the tail probability of the generated null distribution. Correlation networks were built to reveal additional serology features significantly associated with the selected features. Serology features significantly (*p* < 0.05, after a Benjamini-Hochberg correction) correlated (Spearman r_s_ >|0.7|) via Spearman correlation were selected as co-correlates. Correlation coefficients were calculated using the ‘correlate’ function in the ‘Corrr’ package (v0.4.3), with *p* values corrected using ‘p.adjust’ from the ‘stats’ package (v4.0.3). Network visualization was performed using ‘ggraph’ (v2.0.5) and ‘igraph’ (v1.2.6) packages, with manual label and node positioning corrections made in Adobe Illustrator (v2020) for improved visualization. The gradient color of edges represents correlation value between the features, represented as nodes. Nodes are colored according to selected status, with grey nodes as selected features and white nodes as co-correlate features. An R notebook for the analysis is available.

#### Single cell RNA sequencing

All scRNA-seq gene expression analysis was performed using the Human Cell Atlas instance of the Galaxy biocomputing framework (https://humancellatlas.usegalaxy.eu (74, 75)) based on the “Reference-based RNA-seq data analysis” (76) and “Pre-processing of Single-Cell RNA data” (77, 78) workflow templates. Paired-end FASTQ reads were aligned to the human genome (hg19) with gene annotations from Ensembl (Homo_sapiens.GRCh37.75.gtf (79)) using Trimmomatic (80) followed by RNAStar (81). After removing multiply mapped reads, the number of reads mapped to each gene was quantified with FeatureCounts (82) using a GTF file groomed with StringTie Merge (83) to annotate genes. Heterogeneity in gene expression was then explored with either Seurat (84) in R (v4.1.0) – including PCA for downstream Louvain clustering and UMAP visualisation – or with DESeq2 and Annotate DESeq2 in Galaxy. Harmony (85) was used to adjust for the possibility of batch effects prior to the clustering/ UMAP analyses. Expression of genes of interest was compared between cluster 4 and other clusters using Wilcoxon Rank Sum Tests with Bonferroni correction for multiplicity of testing (individual genes; padj). Enrichment of genes in a plasma cell gene set (“TARTE_PLASMA_CELLS_VS_B_LYMPHOCYTE_UP” (32) accessed from https://www.gsea-msigdb.org/gsea/msigdb) was likewise compared with a Kruskal-Wallis test (gene set; *p* value). Differentially expressed genes identified with DESeq2 are reported alongside adjusted *p* values (padj) following an FDR correction (Benjamini-Hochberg).

Pathway analysis was also performed using Fgsea (fast gene set enrichment analysis; Korotkevitch *et al.* bioRxiv 060012; doi: https://doi.org/10.1101/060012) using either all transcript counts, or only those with significantly different expression between groups (padj<0.05 following DESeq2). Hallmark and Kegg mSigDB gene sets were used to define enriched gene sets from previously curated databases. Reproducible Galaxy workflows are available from the Lead Contact on request.

BCR CDR3 repertoire analysis was performed with the MiXCR Analyze shotgun pipeline, designed for clonotype analysis of non-(VDJ)-enriched RNA sequencing data (86, 87). Only productive rearrangements were considered for downstream analysis. If multiple heavy or light chain clones were reported for a given cell the clones with the highest read counts were used for analysis. CDR3 percentage germline mutations were calculated as averages from V-D-J (heavy chain clones only) or V-J only (heavy and light chain clones). CDR3 length and V gene usage are direct outputs of MiXCR. Hierarchical clustering of CDR3 amino acid sequences was performed using Geneious Tree Builder (Alignment type = Global alignment; Genetic Distance Model: Jukes-Cantor; Tree Build Method: Neighbor-Joining). Dendrograms were generated in Geneious (showing unrooted tree as rooted; proportional transformation).

## Additional Resources

Further information on the relevant clinical trial (NCT02927145) can be found here: https://clinicaltrials.gov/ct2/show/NCT02927145.

## Data Availability

Further information and requests for resources and reagents should be directed to and will be fulfilled by the Lead Contact, Carolyn Nielsen.
The scRNA-seq data generated during this study are available from the NCBI SRA database, BioProject accession number SUB10783850.

## Acknowledgments

We thank the volunteers and clinical staff for participating in and running the clinical trials essential for this study, especially Fay Nugent, Yrene Themistocleous, Alison Lawrie, and Ian Poulton. We also acknowledge technical laboratory support from David Ambrose, Andrew Worth and Julie Furze, sequencing analysis advice from Jennifer Hillman-Jackson and Björn Grüning. Finally, assistance with pilot work from Sarah Hamilton, and Marc Lievens, Danielle Morelle and GSK for participation in the study design and supply of the AS01_B_ adjuvant. C.M.N. was supported by a Sir Henry Wellcome Postdoctoral Fellowship (209200/Z/17/Z). S.J.D. was supported by a Wellcome Trust Senior Fellowship (106917/Z/15/Z). S.J.D. was also a Lister Institute Research Prize Fellow and a Jenner Investigator. The ChAd63-MVA trial (NCT02181088) was supported by funding from the European Union Seventh Framework Programme (FP7/2007-2013) under the grant agreement for MultiMalVax (no. 305282). The protein/AS01_B_ trial (NCT02927145) was funded by MRC grant MR/K025554/1 and the Office of Infectious Diseases, Bureau for Global Health, US Agency for International Development (USAID), under the terms of the Malaria Vaccine Development Program (MVDP) contract AID-OAA-C-15-00071, for which Leidos is the prime contractor. Both clinical studies were also supported in part by UK NIHR infrastructure through the NIHR Oxford Biomedical Research Centre. The Freiburg Galaxy Team at the University of Freiburg (Germany) is funded by the Collaborative Research Centre 992 Medical Epigenetics (DFG grant SFB 992/1 2012) and the German Federal Ministry of Education and Research BMBF grant 031 A538A de.NBI-RBC. The Galaxy server is in part funded by Collaborative Research Centre 992 Medical Epigenetics (DFG grant SFB 992/1 2012) and German Federal Ministry of Education and Research (BMBF grants 031 A538A/A538C RBC, 031L0101B/031L0101C de.NBI-epi, 031L0106 de.STAIR (de.NBI)).The opinions expressed herein are those of the authors and do not necessarily reflect the views of the US Agency for International Development. GlaxoSmithKline Biologicals SA was provided the opportunity to review a preliminary version of this manuscript for factual accuracy, but the authors are solely responsible for final content and interpretation.

## Author contributions

C.M.N. led the study. R.O.P., A.M.M., and S.J.D. were chief, principal, or lead investigators on the clinical trials. C.M.N., J.R.B, J.K.F., A.R.M., F.L., and S.E.S. performed experiments. C.M.N, J.R.B., C.D., J.K.F., C. Goh, C. Griffin, A.K., C.L., S.D., M.T., D.A.L., and G.A. analysed and/or reviewed data. F.L., J.F. and A.R. supported project management and training. R.O.P, A.M.M, R.A.S., D.C.D., G.A., and S.J.D. contributed reagents, materials and/or analysis tools. C.M.N. wrote the manuscript.

