## Supplemental Data and Methods for "Delayed booster dosing improves human antigen-specific Ig and B cell responses to the RH5.1/AS01_B_ malaria vaccine"

### Supplemental Figures and Tables

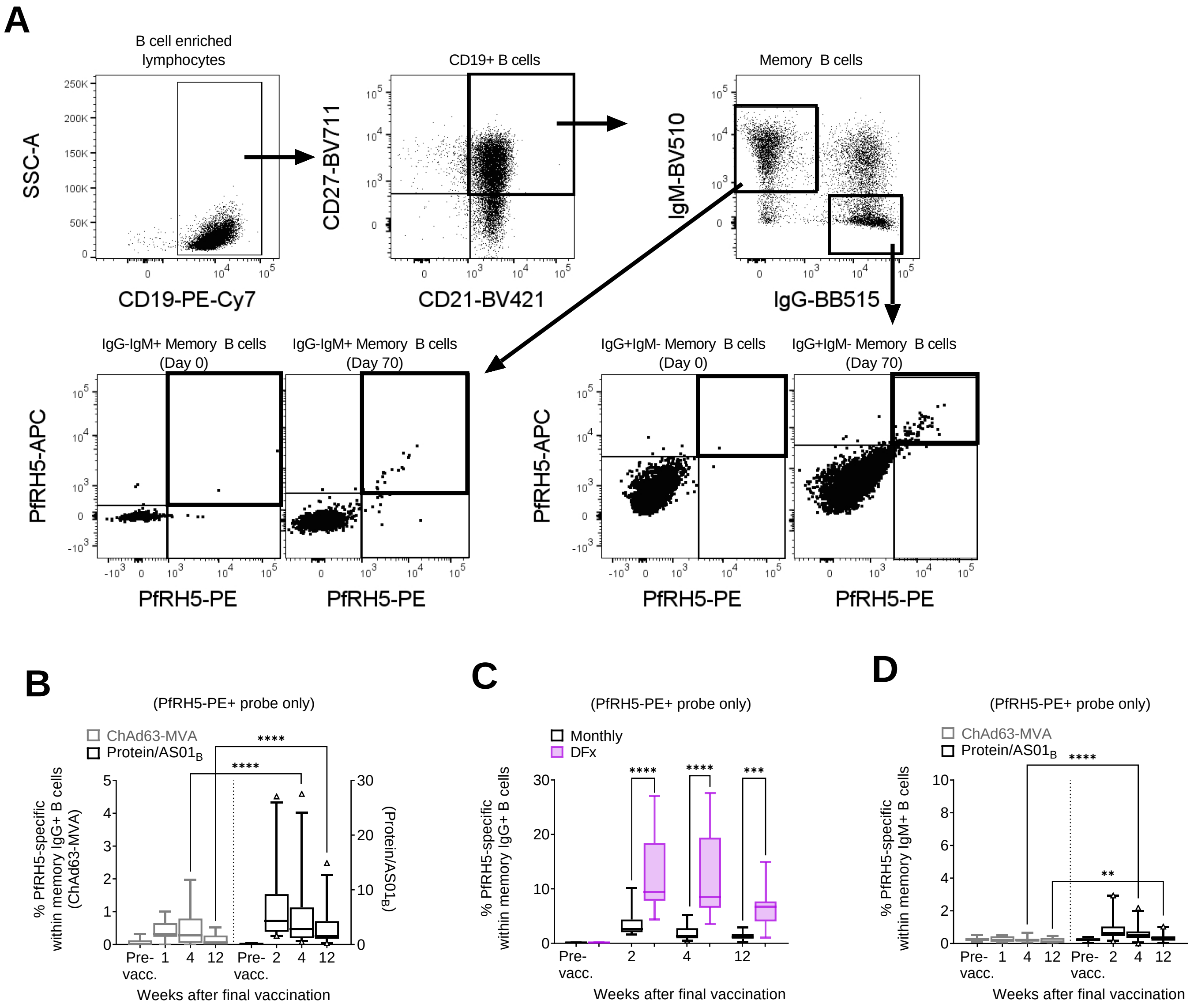
A
B cell enriched lymphocytes
SSC-A
CD19-PE-Cy7
CD19+ B cells
CD27-BV711
CD21-BV421
Memory B cells
IgM-BV510
IgG-BB515
IgG-IgM+ Memory B cells (Day 0)
IgG-IgM+ Memory B cells (Day 70)
IgG+IgM- Memory B cells (Day 0)
IgG+IgM- Memory B cells (Day 70)
PfRH5-APC
PfRH5-PE
B
(PfRH5-PE+ probe only)
ChAd63-MVA
Protein/AS01B
% PfRH5-specific within memory IgG+ B cells (ChAd63-MVA)
(Protein/AS01B)
Pre-vacc.
Weeks after final vaccination
C
(PfRH5-PE+ probe only)
Monthly
DFx
% PfRH5-specific within memory IgG+ B cells
Weeks after final vaccination
D
(PfRH5-PE+ probe only)
ChAd63-MVA
Protein/AS01B
% PfRH5-specific within memory IgM+ B cells
Weeks after final vaccination



2

#### Supplemental Figure 1. Gating strategies for defining PfRH5-specific B cells in flow cytometry-based assays.

**Relates to Figures 1 and 4.**

PBMC from pre-vaccination (Pre-vacc.), and 1-, 2-, 4-, and 12-weeks post final vaccination time points were enriched for B cells and then stained with phenotypic markers and analysed by flow cytometry. (**A**) Gating strategy for PfRH5-specific IgG/ IgM CD19+ B cells within the live lymphocyte population. For data in **Figure 1**, the panel included CD21/CD27 to define memory IgG+ B cells as CD19+CD21+CD27+IgG+IgM-. A more limited panel was used for the single cell sorting as shown in **Figure 4**, to gate the wider CD19+ IgG+ B cell population (no CD21/CD27/IgM staining). For both **Figure 1** and **Figure 4**, PfRH5-specific cells were defined by co-staining with monobiotinylated-PfRH5 conjugated to streptavidin-PE and monobiotinylated-PfRH5 conjugated to streptavidin-APC (PfRH5-PE+PfRH5-APC+). Panels (**B-D**) show the frequencies of PfRH5-specific mBCs when defined only on the basis of PfRH5-PE+. With this less stringent approach, frequencies of PfRH5-specific memory IgG+ B cells (CD19+CD21+CD27+IgG+IgM-) were compared between (**B**) samples from a heterologous viral vector trial (ChAd63-MVA; ChAd63-PfRH5 prime, MVA-PfRH5 boost (10, 11)) and the protein/$AS01_B$ trial, or (**C**) between monthly regimen vaccinees and DFx vaccinees within the protein/$AS01_B$ trial. Frequencies of PfRH5-specific memory IgM+ B cells (CD19+CD21+CD27+IgG-IgM+) were also compared using the PfRH5-PE probe only (**D**). (**B, D**) ChAd63-MVA/ protein/$AS01_B$: Pre-vacc. $n$ = 15/18; 1-week post final vaccination $n$ = 10/0; 2-week $n$ = 0/25; 4-week $n$ = 15/29; 12-week $n$ = 13/25. (**C**) Monthly / DFx within protein/AS01B trial: Pre-vacc. $n$ = 15/3; 2-week $n$ = 16/9; 4-week $n$ = 19/10; 12-week $n$ = 17/8. Comparisons were performed by Mann-Whitney tests. Whiskers denote 5$^{th}$ and 95$^{th}$ percentiles. * $p < 0.05$, ** $p < 0.01$, *** $p < 0.001$, **** $p < 0.0001$.

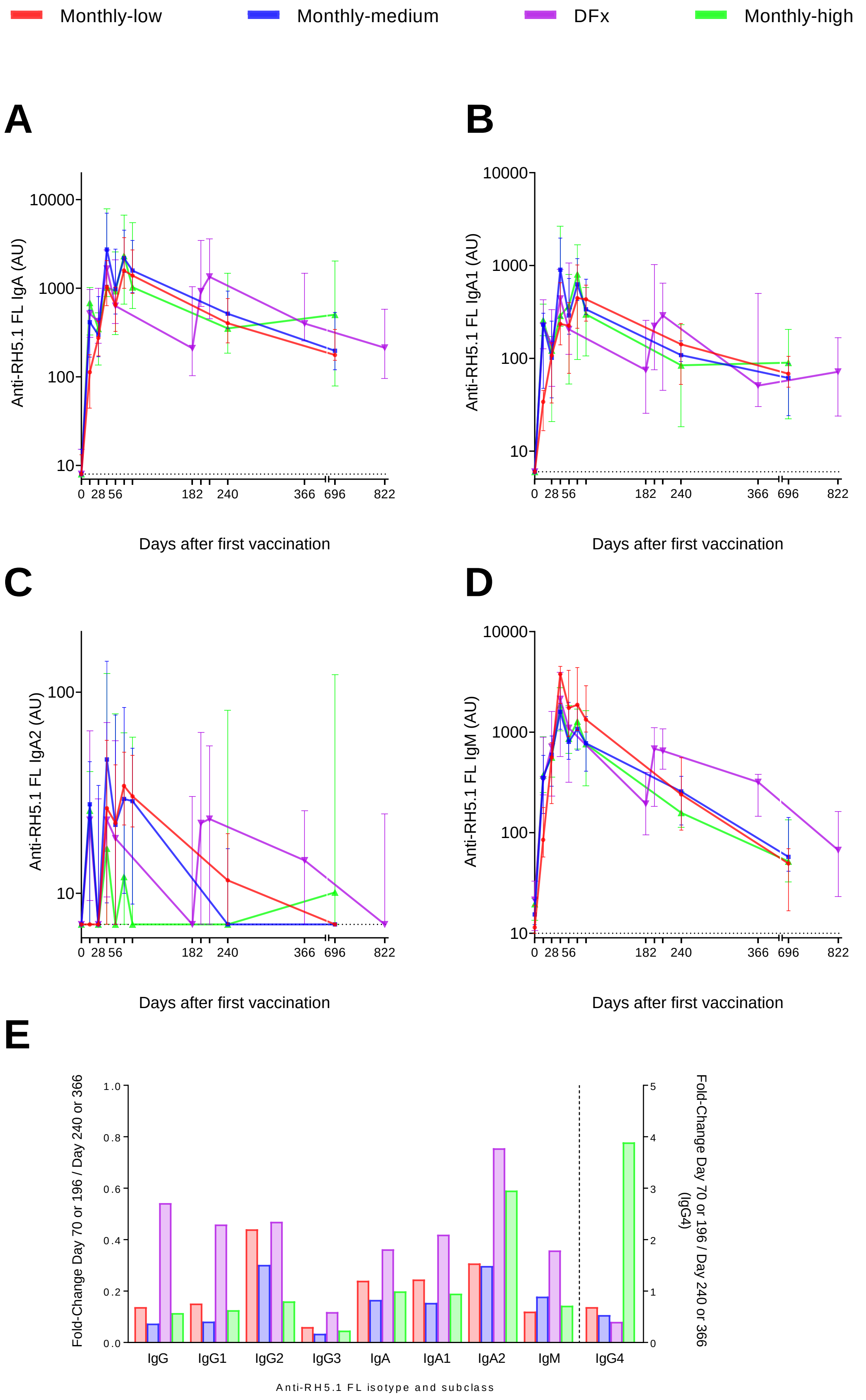
Monthly-low
Monthly-medium
DFx
Monthly-high
A
Anti-RH5.1 FL IgA (AU)
10000
1000
100
10
0 28 56 182 240 366 696 822
Days after first vaccination
B
Anti-RH5.1 FL IgA1 (AU)
10000
1000
100
10
0 28 56 182 240 366 696 822
Days after first vaccination
C
Anti-RH5.1 FL IgA2 (AU)
100
10
0 28 56 182 240 366 696 822
Days after first vaccination
D
Anti-RH5.1 FL IgM (AU)
10000
1000
100
10
0 28 56 182 240 366 696 822
Days after first vaccination
E
Fold-Change Day 70 or 196 / Day 240 or 366
1.0
0.8
0.6
0.4
0.2
0.0
Fold-Change Day 70 or 196 / Day 240 or 366 (IgG4)
5
4
3
2
1
0
IgG
IgG1
IgG2
IgG3
IgA
IgA1
IgA2
IgM
IgG4
Anti-RH5.1 FL isotype and subclass

20

#### Supplemental Figure 2. Antigen-specific IgA and IgM post-vaccination kinetics in DFx and monthly dosing regimens.

**Relates to Figure 1.**

Serum anti-PfRH5.1 FL (full length RH5.1) Ig was assayed by standardised ELISA to report (**A**) total IgA, (**B**) IgGA1 (**C**) IgA2, and (**D**) IgM at key time points (Days 0, 14, 28, 42, 56/182, 70/196, 84/210, 240/366, 696/822). (**E**) Fold change in anti-PfRH5.1 FL Ig between 2-weeks after final vaccination (Day 70 for monthly regimen vaccinees, and Day 196 for DFx vaccinees) and 6-months after final vaccination (Day 240, Day 366). Sample sizes for these ELISAs varied by group and by time point. Monthly-low: $n$ = 12, except Day 696 where $n$ = 9. Monthly-medium: $n$ = 12, except for Day 240 and Day 696 where $n$ = 11 and $n$ = 10, respectively. DFx: $n$ = 12, except for Day 366 and Day 822 where $n$ = 11 and $n$ = 7, respectively. Monthly-high: $n$ = 11, except for Days 70, 240 and 696 where $n$ = 9, $n$ = 10 and $n$ = 4, respectively. Graphs show medians and interquartile ranges.

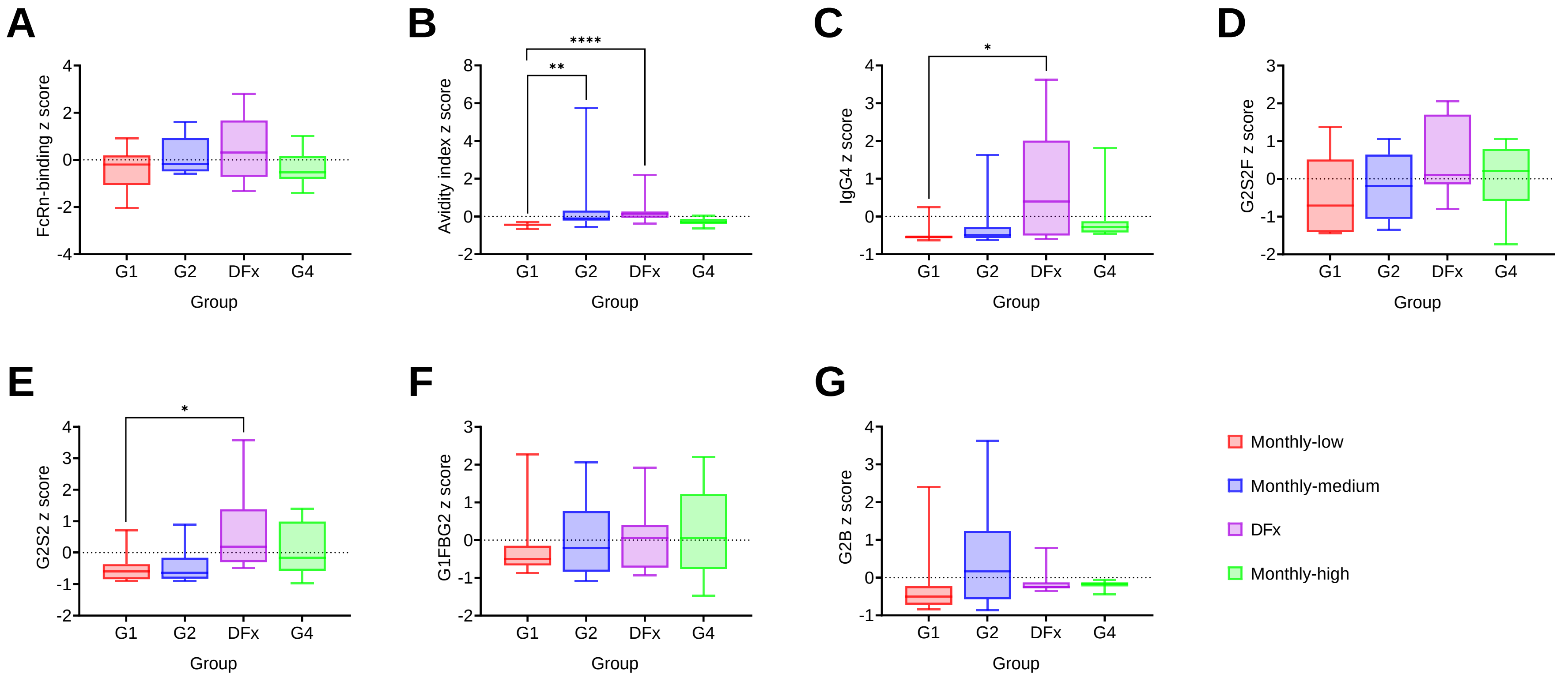


Supplemental Figure 3. Transformed univariate systems serology parameters associated with DFx in computational modelling.

**Relates to Figure 3.**

Systems serology parameters used for computational modelling were first mean-centred and variance-scaled to give Z-scored values. Seven features elevated in DFx vaccinees were subsequently identified as part of larger feature sets discriminating DFx vaccinees from monthly regimen vaccinees, or 'high' dose DFx and monthly-high vaccinees from monthly-low/ monthly-medium vaccinees: FcRn-binding (**A**), avidity index (**B**), IgG4 (**C**), G2S2F (**D**), G2S2 (**E**), G1FBG2 (**F**), and G2B (**G**). Monthly-low: *n* = 12. Monthly-medium: *n* = 11. DFx: *n* = 12, Monthly-high: *n* = 9. Univariate differences were assessed with Kruskal-Wallis tests with Dunn's correction for multiple comparisons. Whiskers denote 5$^{th}$ and 95$^{th}$ percentiles. * $p < 0.05$, ** $p < 0.01$, **** $p < 0.0001$.

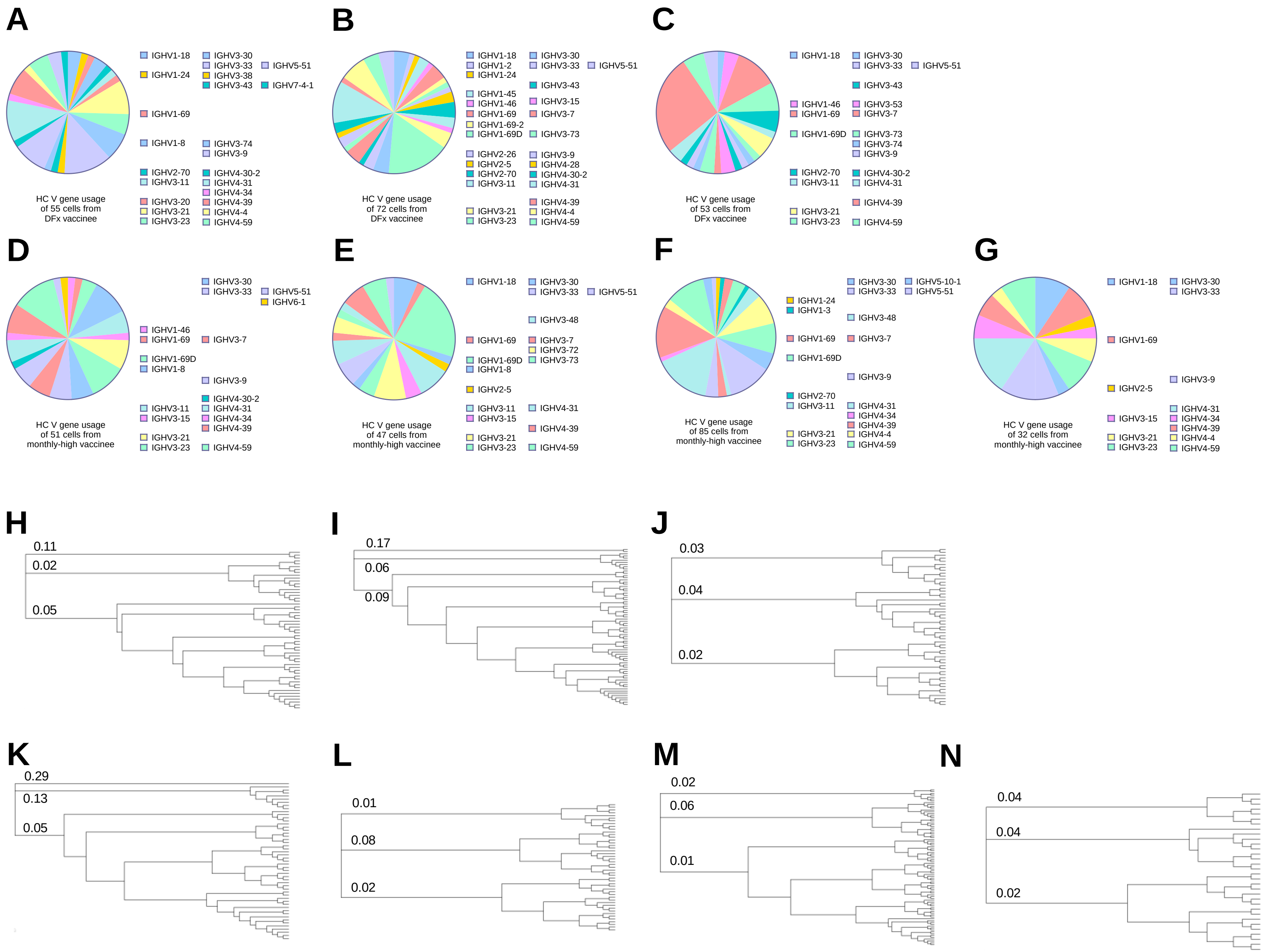

A
HC V gene usage of 55 cells from DFx vaccinee
IGHV1-18
IGHV1-24
IGHV1-69
IGHV1-8
IGHV2-70
IGHV3-11
IGHV3-20
IGHV3-21
IGHV3-23
IGHV3-30
IGHV3-33
IGHV3-38
IGHV3-43
IGHV3-74
IGHV3-9
IGHV4-30-2
IGHV4-31
IGHV4-34
IGHV4-39
IGHV4-4
IGHV4-59
IGHV5-51
IGHV7-4-1
B
HC V gene usage of 72 cells from DFx vaccinee
IGHV1-18
IGHV1-2
IGHV1-24
IGHV1-45
IGHV1-46
IGHV1-69
IGHV1-69-2
IGHV1-69D
IGHV2-26
IGHV2-5
IGHV2-70
IGHV3-11
IGHV3-21
IGHV3-23
IGHV3-30
IGHV3-33
IGHV3-43
IGHV3-15
IGHV3-7
IGHV3-73
IGHV3-9
IGHV4-28
IGHV4-30-2
IGHV4-31
IGHV4-39
IGHV4-4
IGHV4-59
IGHV5-51
C
HC V gene usage of 53 cells from DFx vaccinee
IGHV1-18
IGHV1-46
IGHV1-69
IGHV1-69D
IGHV2-70
IGHV3-11
IGHV3-21
IGHV3-23
IGHV3-30
IGHV3-33
IGHV3-43
IGHV3-53
IGHV3-7
IGHV3-73
IGHV3-74
IGHV3-9
IGHV4-30-2
IGHV4-31
IGHV4-39
IGHV4-59
IGHV5-51
D
HC V gene usage of 51 cells from monthly-high vaccinee
IGHV1-46
IGHV1-69
IGHV1-69D
IGHV1-8
IGHV3-11
IGHV3-15
IGHV3-21
IGHV3-23
IGHV3-30
IGHV3-33
IGHV3-7
IGHV3-9
IGHV4-30-2
IGHV4-31
IGHV4-34
IGHV4-39
IGHV4-59
IGHV5-51
IGHV6-1
E
HC V gene usage of 47 cells from monthly-high vaccinee
IGHV1-18
IGHV1-69
IGHV1-69D
IGHV1-8
IGHV2-5
IGHV3-11
IGHV3-15
IGHV3-21
IGHV3-23
IGHV3-30
IGHV3-33
IGHV3-48
IGHV3-7
IGHV3-72
IGHV3-73
IGHV4-31
IGHV4-39
IGHV4-59
IGHV5-51
F
HC V gene usage of 85 cells from monthly-high vaccinee
IGHV1-24
IGHV1-3
IGHV1-69
IGHV1-69D
IGHV2-70
IGHV3-11
IGHV3-21
IGHV3-23
IGHV3-30
IGHV3-33
IGHV3-48
IGHV3-7
IGHV3-9
IGHV4-31
IGHV4-34
IGHV4-39
IGHV4-4
IGHV4-59
IGHV5-10-1
IGHV5-51
G
HC V gene usage of 32 cells from monthly-high vaccinee
IGHV1-18
IGHV1-69
IGHV2-5
IGHV3-15
IGHV3-21
IGHV3-23
IGHV3-30
IGHV3-33
IGHV3-9
IGHV4-31
IGHV4-34
IGHV4-39
IGHV4-4
IGHV4-59
H
0.11
0.02
0.05
I
0.17
0.06
0.09
J
0.03
0.04
0.02
K
0.29
0.13
0.05
L
0.01
0.08
0.02
M
0.02
0.06
0.01
N
0.04
0.04
0.02

#### Supplemental Figure 4. CDR3 VH gene usage and hierarchical clustering by vaccinee.

**Relates to Figure 5.**

PBMC from pre-vaccination (Pre-vacc.), and 2-weeks post final vaccination in DFx vaccinees (*n*=3) and monthly-high vaccinees (*n*=4) were enriched for B cells and then stained with phenotypic markers for single cell sorting of antigen-specific B cells defined as: live CD19+IgG+ lymphocytes that co-stained for monobiotinylated-PfRH5-PE and monobiotinylated-PfRH5-APC (gating strategy shown in **Supplemental Figure 1A**). Libraries were sequenced following a Smart-Seq v4 and Nextera XT pipeline on a HiSeq4000. CDR3 sequences were extracted using the MiXCR pipeline including heavy chain V gene (VH) usage for DFx (**A-C**) and monthly-high vaccinees (**D-G**) by vaccinee. VH genes in >8% clones per vaccinee are annotated on the pie charts. Hierarchical clustering was performed on CDR3 HC amino acid sequences and visualised as dendrograms for individual monthly-high vaccinees (**H-J**) and DFx vaccinees (**K-N**) by vaccinee. Vaccinees for VH gene usage pie charts are organised in same order as dendrograms, i.e. A=H, B=I, etc. The first three branches are labelled in each dendrogram with branch length (distance between internal nodes) i.e. substitutions per amino acid.

| Cluster | Gene | Average log2(FC) | P-adj |
|---|---|---|---|
| 0 | MS4A1 | 1.227329 | 5.11E-27 |
| | YPEL5 | 1.107417 | 5.07E-20 |
| | CD74 | 1.065549 | 4.59E-26 |
| | DUSP2 | 1.040367 | 1.58E-05 |
| | UCP2 | 1.017806 | 3.14E-14 |
| 1 | *KNG1* | *1.085317* | *1.00E+00* |
| | GAREML | 0.5433 | 1.84E-26 |
| | PSD | 0.531995 | 1.57E-18 |
| | *RP11-217O12.1* | *0.523609* | *1.00E+00* |
| | AL162497.1 | 0.467258 | 1.40E-28 |
| | IRS2 | 0.467258 | 1.40E-28 |
| 2 | LDHA | 1.351499 | 9.71E-09 |
| | CCR7 | 1.348396 | 6.09E-09 |
| | NPM1 | 1.326971 | 2.50E-13 |
| | MIR155HG | 1.303306 | 8.36E-11 |
| | ENO1 | 1.292817 | 1.43E-12 |
| | ENO1-IT1 | 1.292817 | 1.43E-12 |
| 3 | MT-RNR2 | 1.330852 | 9.94E-05 |
| | MUC3A | 1.119706 | 9.63E-07 |
| | *MT-RNR1* | *1.079884* | *1.00E+00* |
| | KCNQ1OT1 | 1.054008 | 3.79E-11 |
| | TAOK1 | 1.037066 | 3.86E-04 |
| | MIR4523 | 1.036084 | 3.61E-4 |
| 4 | *IGLV2-8* | *6.83988* | *7.50E-01* |
| | *MIR650* | *6.83988* | *7.50E-01* |
| | IGHG2 | 6.128872 | 1.11E-10 |
| | RP11-731F5.2 | 6.128872 | 1.11E-10 |
| | IGLC1 | 5.478077 | 1.26E-05 |
| | IGLJ1 | 5.478077 | 1.26E-05 |
| | IGLL5 | 5.478077 | 1.26E-05 |

#### Supplemental Table 1. Top 5 differentially expressed antigen-specific B cell genes per Harmonised UMAP cluster.

**Relates to Figure 4.**

Top five genes shown per UMAP cluster ranked on fold change. Italics denote genes that are not statistically significant following *p* value adjustment. Average log2(FC): average log fold change between cluster and other clusters; padj: adjusted *p* value after correction for multiplicity of testing.

| Pathway | padj | NES | leadingEdge |
|---|---|---|---|
| KEGG_SYSTEMIC_LUPUS_ERYTHEMATOSUS | 0.0313 | 2.11 | HLA-DQA2, HLA-DRA, HLA-DQA1, HLA-DQB1, HLA-DOA, **FCGR2A**, **C7**, HLA-DRB5 |
| KEGG_INTESTINAL_IMMUNE_NETWORK_FOR_IGA_PRODUCTION | 0.0313 | 2.10 | HLA-DQA2, HLA-DRA, HLA-DQA1, HLA-DQB1, HLA-DOA, **IL15**, HLA-DRB5, **TNFRSF17, ITGA4** |
| KEGG_TYPE_I_DIABETES_MELLITUS | 0.0313 | 2.08 | HLA-DQA2, HLA-DRA, HLA-DQA1, HLA-DQB1, HLA-DOA, HLA-C, HSPD1, HLA-DRB5, **IL1A**, HLA-B |
| KEGG_AUTOIMMUNE_THYROID_DISEASE | 0.0313 | 2.01 | HLA-DQA2, HLA-DRA, HLA-DQA1, HLA-DQB1, HLA-DOA, HLA-C, **CGA**, HLA-DRB5, HLA-B, HLA-DOB |
| KEGG_GRAFT_VERSUS_HOST_DISEASE | 0.0464 | 1.97 | HLA-DQA2, HLA-DRA, HLA-DQA1, HLA-DQB1, HLA-DOA, HLA-C, HLA-DRB5, **IL1A,** HLA-B |
| KEGG_PROTEIN_EXPORT | 0.0313 | 1.95 | **SEC11C, SPCS2, HSPA5, SRP68, SEC63, SEC61A1, SPCS1, SEC61G, SPCS3, SEC61A2, SEC61B, SRPRB, SRP54** |
| KEGG_ASTHMA | 0.0464 | 1.95 | HLA-DQA2, HLA-DRA, HLA-DQA1, HLA-DQB1, HLA-DOA, HLA-DRB5 |
| KEGG_ALLOGRAFT_REJECTION | 0.0313 | 1.94 | HLA-DQA2, HLA-DRA, HLA-DQA1, HLA-DQB1, HLA-DOA, HLA-C, HLA-DRB5, HLA-B, HLA-DOB |
| KEGG_CELL_ADHESION_MOLECULES_CAMS | 0.0313 | 1.92 | HLA-DQA2, HLA-DRA, **SELPLG**, HLA-DQA1, HLA-DQB1, HLA-DOA, HLA-C, **SELL**, **ICAM2**, HLA-DRB5, **ITGA4**, HLA-B, HLA-DOB, **PTPRC**, **ITGB1** |
| KEGG_LEISHMANIA_INFECTION | 0.0464 | 1.87 | HLA-DQA2, HLA-DRA, HLA-DQA1, HLA-DQB1, HLA-DOA, **FCGR2A**, HLA-DRB5, **IL1A, ITGA4, NCF1, TLR4,** HLA-DOB, **ITGB1** |

#### Supplemental Table 2. KEGG pathways enriched in monthly-high as compared to DFx dosing vaccinees.

**Relates to Table 3.**

KEGG gene set enrichment analyses were run with the 5,115 significant genes from DESeq2 analyses comparing gene expression in PfRH5-specific CD19+IgG+ B cells from monthly and DFx regimen vaccinees (top thirty genes shown in **Table 2**). All gene set pathways with significant adjusted *p* value (following Benjamini-Hochberg correction for multiplicity of testing) are shown, in order of enrichment. Non-HLA genes are highlighted and underlined. padj: adjusted *p* value; NES: normalised enrichment score; Leading Edge: subset of genes from gene set contributing most to the enrichment signal.

| Pathway | p value | padj | NES | Leading Edge |
|---|---|---|---|---|
| HALLMARK_E2F_TARGETS | 3.03E-03 | 6.28E-02 | 1.62 | PAICS, RFC3, TMPO, HMGB2, CKS1B, RFC1, PTTG1, AK2, XPO1, RAD50, RACGAP1, SMC3, KPNA2, ORC6, BIRC5, TUBG1, EXOSC8, KIF2C, RAD51C, NUDT21, RAD51AP1, CCNB2, EIF2S1, CDKN3, CSE1L, ANP32E, RPA2, ORC2, CDC20, TP53, STMN1, DUT, EED, DONSON, POLD2, NBN, CDK4, PRDX4, NME1, PCNA, XRCC6, TIMELESS, BRCA1, CHEK2, MAD2L1 |
| HALLMARK_OXIDATIVE_PHOSPHORYLATION | 2.01E-03 | 6.28E-02 | 1.61 | PRDX3, MRPL35, MRPL15, UQCRQ, NNT, LDHB, LDHA, MTRF1, NDUFB7, NDUFB4, SLC25A4, SURF1, ECHS1, MRPS11, SDHC, SDHD, PDHB, ATP6V0E1, NDUFB1, ATP6V0B, TIMM8B, TIMM17A, COX15, MRPS12, SLC25A5, NDUFAB1, NDUFB6, FXN, UQCRFS1, UQCRH, COX7A2L, DLAT, CYC1, COX6C, MPC1, UQCRC2, ISCU, NDUFA6, ETFA, CASP7, ATP6V1E1, NDUFV2, MDH1, CYCS, COX6A1, NDUFS6, PDHA1, SDHB, NDUFS1, UQCRC1, ATP6V1G1, MRPL34, HSD17B10, ACO2, DECR1, ATP6V1H, POLR2F, UQCRB, COX5B, PHB2, MDH2, HSPA9, GOT2, AIFM1, MRPL11, TOMM22, ACADVL, GPI |
| HALLMARK_INTERFERON_ALPHA_RESPONSE | 1.68E-02 | 1.68E-01 | 1.61 | SAMD9L, TRIM5, HLA-C, CASP1, SELL, IL15, BST2, PSMB8, PSMA3, PSME1, PSME2, ISG20, UBE2L6, TAP1, B2M, EPSTI1, CD47, TXNIP, IFITM2, MX1, TRAFD1, LAP3, PARP14, CCRL2, SP110, ADAR, CD74, IFITM3, SLC25A28, IRF2, ELF1, IRF1, NCOA7, TDRD7, RNF31 |
| HALLMARK_MTORC1_SIGNALING | 5.03E-03 | 6.28E-02 | 1.58 | SDF2L1, TPI1, GLRX, PSAT1, SHMT2, P4HA1, SLC7A11, HSPA5, XBP1, GAPDH, HSPD1, PPA1, PSMA3, USO1, LDHA, LTA4H, EDEM1, RAB1A, TUBG1, PSMC2, M6PR, SSR1, PDK1, HSP90B1, UFM1, EBP, CACYBP, CANX, ENO1, EIF2S2, ADD3, RPN1, BTG2, SKAP2, ALDOA, TES, BUB1, CYB5B, SLA, SEC11A, PSMC6, SC5D, LGMN, PGM1, SERP1, PSMD12, PGK1, BHLHE40, NUFIP1, PSMD13, FKBP2, TCEA1, GMPS, HSPE1, TUBA4A, CD9, ACTR3, PSMA4, PSMD14, CORO1A, NMT1, HSPA4, ITGB2, PSMB5, HSPA9, PSPH, CCT6A, CALR, YKT6, STIP1, GPI, ETF1, TFRC, ASNS, UCHL5, PHGDH, HPRT1, PSMC4, CCNG1, HMGCR |
| HALLMARK_MYC_TARGETS_V1 | 4.02E-03 | 6.28E-02 | 1.56 | PHB, PRDX3, HSPD1, LDHA, XPO1, TCP1, KPNA2, IMPDH2, CCT5, PSMD7, RRM1, CANX, EIF2S2, SRM, VBP1, SSBP1, CCT3, EIF2S1, NHP2, CDK2, ORC2, CDC20, APEX1, SF3A1, NDUFAB1, DUT, PSMC6, PSMB2, NPM1, POLD2, HNRNPC, CDK4, SF3B3, GNL3, RFC4, CYC1, PRDX4, PGK1, NME1, KPNB1, PTGES3, BUB3, PCNA, EIF3J, HNRNPA2B1, HSPE1, XRCC6, SSB, RNPS1, PSMA4, MAD2L1, PSMD14, PSMD1, CCT7, RPLP0, HNRNPD, TARDBP, SNRPD2, SMARCC1, EIF4H, PHB2, PSMA7, CCT2, GOT2, CNBP, PPM1G, FBL, ETF1, RPS2, CBX3, HPRT1, HNRNPR, PSMC4, HSP90AB1, MRPL23, TUFM, SNRPD3, PSMD8 |
| HALLMARK_UNFOLDED_PROTEIN_RESPONSE | 2.97E-02 | 2.48E-01 | 1.50 | PSAT1, CKS1B, HSPA5, FKBP14, XBP1, EXOSC1, DNAJC3, DNAJB9, SPCS1, EDEM1, HERPUD1, SPCS3, SSR1, PDIA6, GOSR2, EIF4EBP1, SRPRB, HSP90B1, TTC37, EIF2S1, NHP2, PDIA5, EIF4A2, ALDH18A1, SEC11A, NPM1, SERP1, WIPI1, EIF4A3, CXXC1, PREB, PAIP1, TSPYL2, YWHAZ, EXOSC5, DCP2, ATF4, HSPA9, CALR, SLC30A5, NFYB |
| HALLMARK_PROTEIN_SECRETION | 4.38E-02 | 3.07E-01 | 1.47 | LMAN1, BET1, SNX2, USO1, SEC24D, VPS45, TMED10, SNAP23, RAB14, M6PR, NAPA, RAB2A, COPB2, GOSR2, AP2B1, STX7, COPE, TMED2, RAB9A, ARCN1, ANP32E, PPT1, TMX1 |

Supplemental Table 3. HALLMARK pathways enriched in monthly-high as compared to DFx dosing vaccinees.

**Relates to Table 3.**

Hallmark gene set enrichment analyses were run with the 5,115 significant genes from DESeq2 analyses comparing gene expression in PfRH5-specific CD19+IgG+ B cells from monthly-high and DFx regimen vaccinees (top twenty genes shown in **Table 2**). All gene set pathways with significant adjusted *p* values (following Benjamini-Hochberg correction for multiplicity of testing) are shown in order of enrichment. padj: adjusted *p* value; NES: normalised enrichment score; Leading Edge: subset of genes from gene set contributing most to the enrichment signal.

### Supplemental Systems Serology Methods

##### *Fluorescent Primary and Secondary Antibodies*

The following fluorescent antibodies were purchased from BD Biosciences: anti-human CD14-APC-Cy7 (557831), anti-human CD56-PE-Cy7 (335791), and anti-human MIP1β-BV421 (562900). Additional fluorescent antibodies were purchased from BioLegend: anti-human CD66b-PacificBlue (305112), anti-human CD3-APC-Cy7 (300426), anti-human CD3-BV785 (300472), anti-human CD107a-BV605 (328634), and ant-human IFNγ-PE (506507). A FITC-conjugated, goat anti-guinea pig complement C3 polyclonal antibody was purchased from MP Biomedical (0855385). PE-conjugated secondary antibodies were purchased from Southern Biotech for the detection of total human IgG (9040-09), IgM (9020-09), IgA1 (9130-09), IgA2 (9140-09), IgG1 (9052-09), IgG2 (9070-09), IgG3 (9210-09), and IgG4 (9200-09).

##### *Antigen Coupling to Fluorescent Beads*

NeutrAvidin-labelled yellow-green (F8776) and red (F8775) fluorescent 1µm microspheres were purchased from Thermo Fisher Scientific. For immune functional assays, $1.8 \times 10^8$ NeutrAvidin-labelled fluorescent microspheres were coupled to 5µg monobiotinylated PfRH5 antigen (exact construct as described above in flow cytometry section and (10)) by co-incubation in PBS/5% BSA (PBSA) overnight at 4°C, then the beads were washed twice with PBSA. Magplex-C microspheres (Luminex Corp) were covalently coupled to streptavidin (016-000-113, Jackson Immunoresearch) using a two-step carbodiimide reaction. Magplex-C beads ($9 \times 10^8$) were washed, resuspended in 100mM $NaH_2PO_4$ (pH6.2), and activated by incubating with 500µg Sulfo-NHS (A39269, Pierce) and 500µg EDC (A35391, Pierce) for 30 min at room temperature (RT). The beads were washed three times with coupling buffer (50mM MES, pH 5.0), then incubated with streptavidin in 500µL of coupling buffer for 2h at RT. The beads were washed with PBS/0.05% Tween-20, incubated overnight at 4°C with 100µg/mL monobiotinylated PfRH5 in PBSA, washed, and stored in PBS/0.05% sodium azide.

##### *THP-1 Monocyte Phagocytosis Assay*

An assay for measuring antibody-dependent THP-1 monocyte / cellular phagocytosis (ADCP) was used as previously described (87). Briefly, 1µm yellow-green fluorescent NeutrAvidin beads were coupled to monobiotinylated PfRH5 antigen and blocked overnight with PBSA. The beads were then washed twice with PBSA, diluted to $1.8 \times 10^8$ beads/mL, and 10µL beads/well were added to a 96-well round-bottom microplate. Diluted plasma from immunised subjects (10µL/well) was added to the beads and incubated at 37°C for 2h to allow the formation of immune complexes. Unbound antibodies were washed off, then 25,000 THP-1 cells/well (TIB202, ATCC) were added to the beads in 200µL THP-1 medium (R10 + 55 mM β-ME) and incubated overnight at 37°C. Cells were fixed and acquired on an Intellicyt iQue Screener PLUS flow cytometer. The phagocytic score for each sample was calculated as (% bead-positive cells) x (geometric median fluorescence intensity [MFI] of bead-positive cells) / (10 x gMFI of first bead-positive peak).

##### *Primary Neutrophil Phagocytosis Assay*

An assay for measuring antibody-dependent neutrophil phagocytosis (ADNP) has been described previously (88). Briefly, 1µm yellow-green fluorescent NeutrAvidin beads were coupled to monobiotinylated PfRH5 antigen and blocked with PBSA overnight at 4°C. The beads were then washed twice with PBSA and diluted to $1.8 \times 10^8$ beads/mL. PfRH5-coupled beads (10µL/well) and diluted test plasma (10µL/well) were combined in a round-bottom 96-well plate, then incubated at 37°C for 2h. Primary leucocytes were isolated from freshly drawn whole blood (collected from healthy donors in anticoagulant citrate dextrose tubes) by treatment with ACK red blood cell lysis buffer, then diluted in R10 media to 250,000 cells/mL. After immune complex formation, the beads were washed, combined with 50,000 primary leucocytes/well, and incubated for 1h at 37°C. Cells were stained for surface CD66b, CD14, and CD3, fixed, and acquired on an Intellicyt iQue Screener PLUS flow cytometer. Gates were drawn to identify singlet $SSC^{hi}$CD66b+CD14-CD3- cells, and phagocytic scores for each sample were calculated as (% bead-positive cells) x (gMFI of bead-positive cells) / (10 x gMFI of the first bead-positive peak).

*Complement Deposition Assay*

An assay for measuring antibody-dependent complement deposition (ADCD) was used as previously described (89). Briefly, 1mm red fluorescent NeutrAvidin beads were incubated with monobiotinylated PfRH5 antigen, blocked with PBSA, then washed and diluted to $1.8x10^8$ beads/mL. PfRH5-coupled beads (10µL/well) were combined with diluted test plasma (10µL/well) in a 96-well round-bottom microplate, then incubated at 37°C for 2h. Guinea pig complement (CL4051, CedarLane) was diluted in gelatin veronal buffer containing calcium and magnesium (GVB++; IBB-300, Boston Bioproducts). The beads were washed with PBS and incubated with diluted complement for 20 min at 37°C. The beads were then washed with 5mM EDTA, stained with FITC-conjugated anti-complement C3, and acquired on an Intellicyt iQue Screener PLUS flow cytometer. Gates were drawn on singlet, red fluorescent particles, and complement deposition was reported as the median FITC fluorescence intensity.

*NK cell Activation Assay*

An assay for measuring antibody-dependent NK cell activation (ADNKA) has been described previously (90, 91). Flat-bottom 96-well ELISA plates (439454, Thermo Fisher) were coated with monobiotinylated PfRH5 antigen, then blocked with PBSA. Plasma samples from test subjects were diluted in PBSA, added to the plates, and incubated for 2h at 37°C. Primary human NK cells were purified from buffy coats from healthy donors using the RosetteSep human NK cell enrichment cocktail (15065, StemCell), then resuspended in R10 media containing 10µg/mL brefeldin A (B7651, Sigma), GolgiStop (554724, BD Biosciences), and fluorescent anti-human CD107a. The ELISA plates were washed three times with PBS, then isolated NK cells (25,000/well) were added and incubated at 37°C for 5h. The cells were then stained for surface CD56 and CD3, permeabilised, stained with fluorescent antibodies to IFNγ and MIP1β, fixed, and acquired on an Intellicyt iQue Screener PLUS flow cytometer. Gates were drawn on singlet, CD56+/CD3- cells, and results were reported as the percentages of these cells that expressed surface CD107a, intracellular MIP1β, or intracellular IFNγ.

*Antibody Isotype and Subclass Analysis*

The isotypes and subclasses of PfRH5 antigen-specific antibodies were quantified using a previously described method (92). Magplex-C microspheres were coupled to streptavidin via carbodiimide crosslinking with Sulfo-NHS and EDC. Streptavidin-coupled beads were then incubated overnight with monobiotinylated PfRH5 antigen, blocked with PBSA, and added to black flat-bottom 384-well plates (781906, Greiner BioOne) so that each well contained 1500 PfRH5-coupled beads. Plasma from test subjects was diluted in PBSA and co-incubated with the beads for 2h at RT on a plate shaker (800rpm). The beads were then washed and incubated with a PE-conjugated antibody to detect total human IgG, huIgG1, huIgG2, huIgG3, huIgG4, huIgM, huIgA1, or huIgA2 for 1h at RT on a plate shaker (800rpm). To evaluate anti-PfRH5 IgG avidity, samples were incubated with or without 7M urea prior to incubation with total human IgG antibody. The beads were then washed and acquired on an Intellicyt iQue Screener PLUS flow cytometer. Results for all 9 parameters were reported as the median PE fluorescence intensity. For the avidity assay, an avidity index is reported as (MFI with urea incubation) / (MFI without urea incubation). To note, previously published data on anti-PfRH5 IgG avidity was generated using a sodium thiocyanate chemical displacement ELISA with post-vaccination sera samples (1).

*Fc-binding Protein Array*

The binding of PfRH5 antigen-specific antibodies to human Fc receptors (FcR) and complement C1q was measured using a previously-described assay (93, 94). Briefly, avitagged FCGR2A-H, FCGR2A-R, FCGR2B, FCGR3A-F, FCGR3A-V, FCGR3B, FcRn, and FcαR proteins were produced and purified by the Duke Human Vaccine Institute Protein Production Facility. These proteins were then biotinylated with BirA ligase using a commercially available kit (BirA500, Avidity). Purified human C1q protein (C1740, Sigma) was biotinylated using EZ-Link Sulfo-NHS-LC-LC-Biotin (A35358, Pierce) according to the manufacturer's instructions. These biotinylated Fc domain-binding proteins were then incubated with streptavidin-PE (PJ31S, Prozyme) to generate the assay detection reagents. Magplex-C microspheres were coupled to monobiotinylated PfRH5 antigen as described above, blocked with PBSA, and added to 384-well plates so that each well contained 1500 PfRH5-coupled beads. Plasma from test subjects was diluted in PBSA, added to the beads, and incubated for 2h at RT on a plate shaker (800rpm). The beads were then washed, incubated with one of the PE/FcR conjugates for 1h at RT on a plate shaker (800rpm), washed again, and acquired on an Intellicyt iQue Screener PLUS flow cytometer. Results for all 9 parameters were reported as the median PE fluorescence intensity.

##### *Fc glycan Analysis*

The Fc glycans on PfRH5 antigen-specific IgG were analysed using a previously described method (95, 96). Briefly, 200µL of plasma from each vaccinated subject was heat-inactivated at 56°C for 1h, then centrifuged at 20,000xg for 10min at RT. The resulting supernatant samples were first pre-cleared by incubating with 1mm magnetic streptavidin-coated microspheres (S1420S, New England Biolabs) for 1h at RT. A magnet was used to pellet the beads, and the supernatants were then transferred to new tubes containing streptavidin-coated beads that had been coupled to monobiotinylated PfRH5 antigen. The PfRH5-coupled beads were incubated with plasma samples for 1h at 37°C, then washed and incubated with IdeZ enzyme (P0770S, New England Biolabs) for 1h at 37°C to remove the Fc fragments from the bead-bound antibodies. These Fc fragments were transferred to new tubes and incubated with PNGase F (A28404, Applied Biosystems) for 1h at 50°C to remove the glycans. The glycans were then isolated and labelled with APTS dye using a GlycanAssure kit (A28676, Thermo Fisher) according to the manufacturer's instructions. Finally, APTS-labelled glycan samples were analysed by capillary electrophoresis on an ABI 3500xL Genetic Analyzer. The area under the peak for each Fc glycan structure was calculated using GlycanAssure data analysis software. Results were then reported as the frequency (%) of each glycan structure within a given sample. A total of 25 glycan structures were measured, of which 11 were detectable in >50% vaccinees and included in downstream analyses: G2S2, G2S2F, G1S1F, G2S1F, G0F, G1F, G1.F-G1FB, G1.FB-G2, G2B, G2F, and G2FB.
